# Association of assisted reproductive technology with offspring growth and adiposity from infancy to early adulthood

**DOI:** 10.1101/2022.03.20.22272579

**Authors:** Ahmed Elhakeem, Amy E. Taylor, Hazel M. Inskip, Jonathan Huang, Muriel Tafflet, Johan L. Vinther, Federica Asta, Jan S. Erkamp, Luigi Gagliardi, Kathrin Guerlich, Jane Halliday, Margreet W. Harskamp-van Ginkel, Jian-Rong He, Vincent WV. Jaddoe, Sharon Lewis, Gillian M. Maher, Yannis Manios, Toby Mansell, Fergus P McCarthy, Sheila W. McDonald, Emanuela Medda, Lorenza Nisticò, Angela Pinot de Moira, Maja Popovic, Irwin KM. Reiss, Carina Rodrigues, Theodosia Salika, Ash Smith, Maria A. Stazi, Caroline Walker, Muci Wu, Bjørn A. Åsvold, Henrique Barros, Sonia Brescianini, David Burgner, Jerry KY. Chan, Marie-Aline Charles, Johan G. Eriksson, Romy Gaillard, Veit Grote, Siri E. HÅberg, Barbara Heude, Berthold Koletzko, Susan Morton, George Moschonis, Deirdre Murray, Desmond O’ Mahony, Daniela Porta, Xiu Qiu, Lorenzo Richiardi, Franca Rusconi, Richard Saffery, Suzanne C. Tough, Tanja GM. Vrijkotte, Scott M. Nelson, Anne-Marie Nybo Andersen, Maria C. Magnus, ART-Health Cohort Collaboration, Deborah A. Lawlor

**Affiliations:** MRC Integrative Epidemiology Unit at the University of Bristol, Bristol, UK; Population Health Science, Bristol Medical School, University of Bristol, Bristol, UK; NIHR Bristol Biomedical Research Centre, Bristol, UK; MRC Lifecourse Epidemiology Centre, University of Southampton, Southampton, UK; NIHR Southampton Biomedical Research Centre, University of Southampton and University Hospital Southampton NHS Foundation Trust, Southampton, UK; Singapore Institute for Clinical Science, Agency for Science, Technology, and Research, Singapore; Université de Paris, Inserm, INRAE, Centre for Research in Epidemiology and StatisticS (CRESS), Paris, France; Section of Epidemiology, Department of Public Health, University of Copenhagen, Copenhagen, Denmark; Depatment of Epidemiology, Lazio Regional Health Service, Rome, Italy; The Generation R Study Group, Erasmus MC, University Medical Center, Rotterdam, The Netherlands; Department of Paediatrics, Erasmus MC, University Medical Center, Rotterdam, The Netherlands; Department of Mother and Child Health, Ospedale Versilia, Viareggio, AUSL Toscana Nord Ovest, Pisa, Italy; LMU - Ludwig Maximilians Universität Munich, Division of Metabolic and Nutritional Medicine, Department of Pediatrics, Dr. von Hauner Children’s Hospital, LMU University Hospitals, Munich, Germany; Murdoch Childrens Research Institute, Parkville, VIC, Australia; University of Melbourne, Parkville, VIC, Australia; Amsterdam UMC, University of Amsterdam, Department of Public and Occupational Health, Amsterdam Public Health Research Institute, Amsterdam, The Netherlands; Division of Birth Cohort Study, Guangzhou Women and Children’s Medical Center, Guangzhou Medical University, Guangzhou, China; School of Public Health, University College Cork, Cork, Ireland; The Irish Centre for Maternal and Child Health Research (INFANT), University College Cork, Cork, Ireland; Department of Nutrition and Dietetics, School of Health Science and Education, Harokopio University, Athens, Greece; Institute of Agri-Food and Life Sciences, Hellenic Mediterranean University Research Centre, Heraklion, Greece; Department of Obstetrics and Gynaecology, University College Cork, Cork, Ireland; Department of Paediatrics, Cumming School of Medicine, University of Calgary, Calgary, Canada; Department of Community Health Sciences, Cumming School of Medicine, University of Calgary, Calgary, Canada; Center for Behavioral Sciences and Mental Health-Istituto Superiore di Sanità, Rome, Italy; Cancer Epidemiology Unit, Department of Medical Sciences, University of Turin and CPO Piemonte, Turin, Italy; EPIUnit - Instituto de Saúde Pública, Universidade do Porto, Porto, Portugal; Laboratório para a Investigação Integrativa e Translacional em Saúde Populacional (ITR), Porto, Portugal; Centre for Longitudinal Research - He Ara ki Mua, Faculty of Medical and Health Sciences, University of Auckland, Auckland, New Zealand; K.G. Jebsen Center for Genetic Epidemiology, Department of Public Health and Nursing, Faculty of Medicine and Health Sciences, NTNU, Norwegian University of Science and Technology, Trondheim, Norway; HUNT Research Centre, Department of Public Health and Nursing, Faculty of Medicine and Health Sciences, NTNU, Norwegian University of Science and Technology, Levanger, Norway; Department of Endocrinology, Clinic of Medicine, St. Olavs Hospital, Trondheim University Hospital, Trondheim, Norway; Department of Paediatrics, University of Melbourne, Parkville, VIC, Australia; Department of Paediatrics, Monash University, Clayton, VIC, Australia; Department of Reproductive Medicine, KK Women’s and Children’s Hospital, Singapore; Academic Clinical Program in Obstetrics and Gynaecology, Duke-NUS Medical School, Singapore; Ined, Inserm, EFS Joint Unit Elfe, Paris, France; Department of Obstetrics and Gynaecology and Human Potential Translational Research Programme, Yong Loo Lin School of Medicine, National University of Singapore, Singapore; Department of General Practice and Primary Health Care, University of Helsinki and Helsinki University Hospital, Helsinki, Finland; Folkhälsan Research Center, Helsinki, Finland; Centre for Fertility and Health, Norwegian Institute of Public Health, Oslo, Norway; Department of Food, Nutrition and Dietetics, School of Allied Health, Human Services and Sport, La Trobe University, Melbourne, Australia; Department of Pediatrics and Child Health, University College Cork, Cork, Ireland; National Longitudinal Study of Children in Ireland, Economic and Social Research Institute, Dublin, Ireland; School of Medicine, University of Glasgow, Glasgow, UK

**Author notes:** <u>Corresponding author:</u> Dr Ahmed Elhakeem, MRC Integrative Epidemiology Unit at the University of Bristol, Bristol, UK; Population Health Science, Bristol Medical School, University of Bristol, Bristol, UK.

**Keywords:** BMI, Cohort, Height, IVF, Weight

## Abstract

**Importance:** People conceived using assisted reproductive technology (ART) make up an increasing proportion of the world’s population, and their numbers are expected to continue rising.

**Objective:** Investigate association of ART conception with growth and adiposity outcomes from infancy to early adulthood in offspring from a large multinational multi-cohort study.

**Design:** 26 population-based cohort studies.

**Setting:** Europe, Asia-Pacific, and North America

**Participants:** Infants, children, adolescents, and young adults born from 1984 to 2018, with mean ages at assessment of growth/adiposity outcomes ranging from 0.6 month to 27.4 years.

**Exposures:** Conception by ART (conventional *in vitro* fertilisation and intracytoplasmic sperm injection) versus natural conception (NC).

**Main Outcomes and Measures:** Length/height, weight, and body mass index (BMI). Each cohort was analysed separately with adjustment for maternal BMI, age, smoking, education, parity, ethnicity, and offspring sex and age. Cohort results were combined in random effects meta-analysis for thirteen age groups.

**Results:** Up to 158,066 offspring (4,329 conceived by ART) were included in each age-group meta-analysis; 47.6% to 60.6% were female. Compared with NC, ART-conceived offspring were slightly shorter, lighter, and thinner from infancy to early adolescence. The differences in growth/adiposity outcomes were largest at the youngest ages and attenuated with older child age, e.g., adjusted standardised mean differences (95% confidence intervals) in offspring weight at age ‘<3 months’, ‘17 to 23 months’, ‘6 to 9 years’, and ‘14 to 17 years’ were -0.27 standard deviation (SD) units (−0.39 to -0.16), -0.16SD (−0.22 to -0.09), -0.07SD (−0.10 to -0.04), and -0.02SD (−0.15 to 0.12), respectively. There was no evidence that results were driven by parental subfertility or of difference between conventional *in vitro* fertilisation and intracytoplasmic sperm injection however, smaller offspring size appeared to be limited to offspring conceived by fresh but not frozen embryo transfer, compared with NC. More marked but less precise differences were observed for body fat measurements. There was imprecise evidence that offspring conceived by ART may develop greater adiposity by early adulthood.

**Conclusions and Relevance:** People conceiving or conceived by ART can be reassured that differences in early growth and adiposity are small and no longer evident by late adolescence.

**KEY POINTS:** *Question:* Is conception by assisted reproductive technology associated with growth and adiposity from infancy to early adulthood?

*Findings:* In this multi-cohort study of up to 158,066 European, Asian-Pacific, and Canadian infants, children, adolescents, and young adults, those conceived using assisted reproductive technology were on average shorter, lighter, and thinner from infancy up to early adolescence when compared with their naturally conceived peers though differences were small across all ages and reduced with older age.

*Meaning:* Parents conceiving or hoping to conceive through assisted reproductive technology and their offspring should be reassured that differences in early life growth and adiposity are small and no longer apparent by late adolescence.

---

Assisted reproductive technology (ART), which mainly involves *in vitro* fertilisation (IVF) and intracytoplasmic sperm injection (ICSI), has resulted in over 8 million births worldwide during the last four decades (1, 2), and use of ART is expected to continue rising for several reasons, including increasingly delayed childbearing (3, 4). Ever since the first ART birth in 1978, the primary research focus has been on improving live-birth rates (5). Now that ART is acknowledged as an effective procedure for infertility treatment, attention has shifted towards identifying and reducing any adverse effects of ART on maternal or offspring health. Studies examining growth-related outcomes have mostly considered perinatal measures, with results showing an increased risk of low birthweight, small-for-gestational-age, and preterm birth in those conceived using ART (6-9). Furthermore, studies comparing ART procedures suggest perinatal differences between conventional IVF and ICSI (10), and between fresh and frozen embryo transfers (11-14).

Besides perinatal outcomes, long-term associations between ART conception and offspring growth and adiposity remain largely unknown, with the few studies that have examined these mostly limited by small sample size, short follow-up, and limited adjustment for confounders or overadjustment for possible mediators (15-17). A recent study which examined trajectories of change in height, weight, and body mass index (BMI) from birth to age 7 years (n=81,461 offspring with 1,721 conceived by ART) in the Norwegian Mother, Father and Child Cohort Study (MoBa) found that ART-conceived offspring started smaller and grew faster than NC offspring (18). Another more recent but considerably smaller Singaporean birth cohort study (n=1,180 offspring with 85 conceived by ART) discovered smaller height and lower skinfold thickness at age 6 years in ART-conceived than NC offspring (19).

Our primary aim was to conduct a multi-cohort study to provide evidence on the associations of ART conception (compared with NC) with offspring growth and adiposity from infancy to early adulthood. We additionally compared results according to parental subfertility status, in males and females, in ICSI and conventional IVF, and in fresh and frozen embryo transfers.

## METHODS

This multi-cohort study was carried out within the newly established Assisted Reproductive Technology and future Health (ART-Health) consortium, following a pre-specified analysis plan (https://osf.io/qhwvc/), and is reported in line with The Strengthening the Reporting of Observational Studies in Epidemiology (STROBE) Statement guidelines (20).

### Cohort studies

Eligible cohorts were identified from the European Union Child Cohort Network (21-23) and by searching cohort profile papers. We targeted population-based cohorts without selection or oversampling of ART-conceived offspring to reduce potential for selection bias and to ensure identical growth and adiposity assessments for ART-conceived and NC offspring. All cohorts from any geographical region with birth years from older to more contemporary cohorts were eligible for inclusion provided they had data on whether offspring were conceived by ART or NC, and one or more offspring growth or adiposity outcome measures assessed from age one month (including repeated measurements). A total of thirty cohorts were invited to participate and 26 were included in this study (**eTable 1**). Detailed description of the 26 included cohorts is provided in the **eMethods**.

All cohorts had ethical approval from the relevant local or national ethics committees and all offspring gave informed consent/assent to participate in the respective cohorts and secondary data analyses. Details on ethics approvals and consent are in **eMethods**.

### Mode of conception and fertility treatment

Fertility treatment use was defined according to the International Glossary on Infertility and Fertility Care (1). Information on mode of conception and fertility treatment was collected using questionnaires or by record linkage (**eMethods**). This information was used to identify if offspring were conceived by ART (i.e., IVF or ICSI) or were naturally conceived (NC) i.e., without any fertility treatment. We additionally identified (i) if ART conception involved IVF or ICSI, (ii) whether fresh embryo transfer (ET) or frozen-thawed embryo transfer (FET) was used, and (iii) whether NC offspring were born to fertile or sub-fertile parents, depending on a time to pregnancy of within 12 months or >12 months after they begun trying, respectively (1, 24).

### Offspring growth/adiposity outcomes

Primary outcomes for this study were length/height (centimetre (cm)), weight (kilograms (kg)), BMI (kg/m^2^), and secondary outcomes were waist circumference (cm), total body fat %, and fat mass index (FMI; kg/m^2^). Length/height and weight were obtained from research clinics, child health records, and maternal-/self-reports, with BMI calculated from these as weight in kilograms divided by height in metres squared. Waist circumference was mostly measured during research clinics. Body fat % was calculated from bioelectrical impedance analysis done during research clinics, and FMI was derived as fat mass in kilograms from dual-energy x-ray absorptiometry scans divided by height in metres squared. Details on outcome measurements and ages in each cohort are in the **eMethods**. Descriptive data on outcomes and ages at outcome assessments are in **eTable 2**.

Outcome age groups for this study were determined by available data from each cohort i.e., ages at outcomes assessment. Cohorts were allocated to meta-analysis age groups by mean age at outcome assessment with the aim of maximising cohort numbers in each age group meta-analysis. The primary outcomes were allocated to thirteen age groups, and secondary outcomes (available in 17/26 cohorts) were allocated to four age groups. If a cohort had >1 outcome assessment in an age group, we selected the one with the biggest sample size.

### Confounders

We used a Directed Acyclic Graph (**eFigure 1**) to identify and adjust for confounders i.e., anything that could plausibly cause ART use and influence offspring growth/adiposity (25, 26). This process identified the following maternal factors as potential confounders: age at pregnancy/birth, socioeconomic position (education), pre/early-pregnancy BMI, pre/early-pregnancy smoking, parity, and ethnicity. Nineteen cohorts were able to adjust for all these confounders, four did not adjust for ethnicity but were ethnically homogeneous, one did not adjust for parity because it only included nulliparous women, and two were unable to adjust for BMI and smoking. Details on the available confounders in each cohort is in **eMethods**.

### Statistical analysis

Analyses were performed separately in each cohort applying standard statistical code, with results combined using meta-analysis. In cohort-specific analyses, we estimated associations of ART conception (versus NC) with offspring outcomes using linear regression adjusted for confounders (plus offspring age and sex). Analysis was done in offspring with data on mode of conception, ≥1 growth/adiposity outcome, and confounders. To facilitate comparison of results for different outcomes and ages, outcomes were analysed in age-, sex- cohort-specific standard deviation (SD) units (mean=0, SD=1). Cohort results were subsequently combined by random-effects meta-analyses in sub-groups defined by mean age at outcome assessment. Variability in the pooled estimates that is due to between-cohort heterogeneity was quantified by the *I*^*2*^ statistic (27). Influential cohorts (i.e., whose exclusion led to considerable change in the meta-analysis model) were identified by repeating each meta-analyses with each cohort left out in turn.

To separate effects of ART from any effects of parental subfertility, we repeated analyses comparing ART-conceived with NC offspring of sub-fertile parents and separately for NC offspring from fertile parents. Differences by sex and ART treatment types were explored by repeating analyses stratified by sex; comparing IVF/ICSI separately with NC; and comparing fresh ET/FET separately with NC. Lastly, we explored if results reflected effects on multiple births by repeating analysis in singletons and investigated if results were mediated by birth size and pregnancy duration by including extra adjustments (on top of confounders) for birth weight and gestational age.

## RESULTS

A total of 26 cohorts with participants from Europe (n=20 cohorts), Australia (n=2 cohorts), and one each from New Zealand, China, Singapore, and Canada was included in this study (**eTable 1**). Most (n=23 cohorts) were population-based cohorts, two were twins register cohorts, and one was a clinical cohort of ART-conceived young adults and NC controls from the same source population. Birth years were from 1984-2018, with most (n=19 cohorts) born >2002. Mean age at outcome ranged from 0.6 month to 27.4 years. Fifteen cohorts included singletons and multiple births (proportion of multiple births across these ranged from 0.9% to 12.9%), nine included singletons only, and two included twins only. Between 3 to 16 cohorts were included in each meta-analysis with numbers of participants in each meta-analysis ranging from 158,066 (4,329 ART) for weight at age 3 to 5 months to 3,111 (151 ART) for FMI at age >17 years.

Mean length/height was on average smaller in ART-conceived than NC offspring across all age groups, although for some ages, point estimates were close to the null value which was included in the CIs (**Figure 1**). The largest differences in length/height were at the youngest ages and these differences attenuated with older child age. ART-conceived offspring were more similar in height to NC older adolescents and young adults although estimates were imprecise (**Figure 1**).

**Figure 1.**
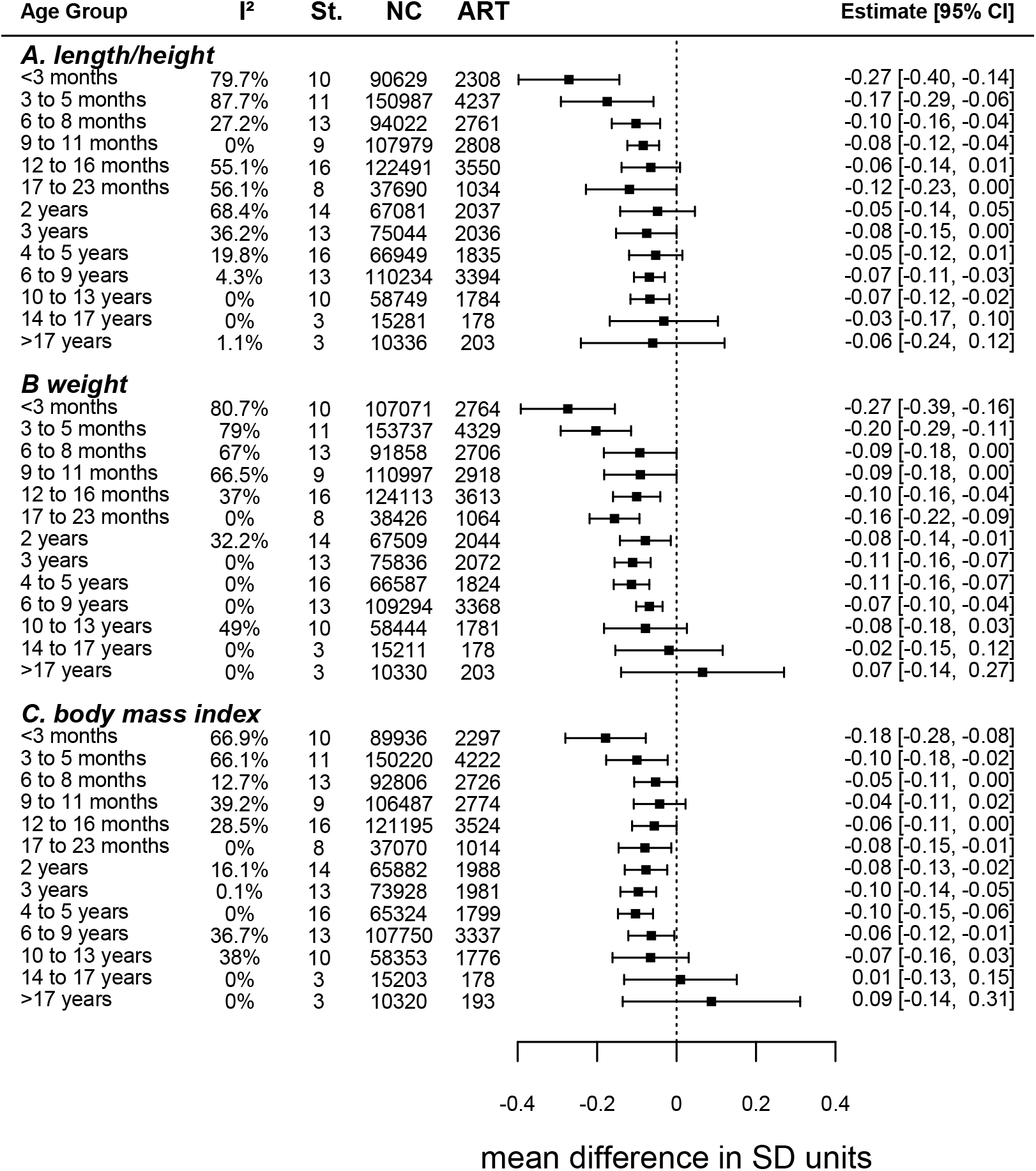
Parts A-C. Mean difference in (A) length/height, (B) weight, and (C) body mass index between ART-conceived and NC offspring. Figure shows the pooled adjusted mean differences in SD units [and 95% confidence intervals] in (A) length/height, (B) weight, and (C) body mass index at each age group between ART-conceived and NC offspring (ART minus NC). Cohort-specific results were adjusted for maternal age, parity, BMI, smoking, education, ethnicity/country of birth, plus offspring sex and age. St. is the number of cohort studies; NC is the number of NC ofspring; ART is the number of ART-conceived offspring; I² represents the percentage of total variability that is due to between cohort heterogeneity. Cohort-specific results are provided in eFigure 2-4.

Mean weight was lower in ART-conceived than NC offspring from age <3 months up to age 10 to 13 years though CIs included the null at ages 6 to 8 months, 9 to 11 months, and 10 to 13 years (**Figure 1**). As for length/height, differences were greatest at the youngest ages and smaller at older offspring ages. The difference in mean weight was close to the null in older adolescents and mean weight in young adults was slightly higher in ART-conceived than NC offspring but with wide CIs (**Figure 1**).

Differences in mean BMI followed a similar pattern to that of weight, with mean BMI lower in ART-conceived than NC offspring up to age 10 to 13 years, with differences being greatest at youngest ages but with wide CIs that included the null value for some results (**Figure 1**). As for weight, difference in mean BMI was closest to the null in older adolescents and mean BMI was somewhat slightly higher for ART-conceived than NC young adults although this was imprecisely estimated (**Figure 1**).

Results for waist circumference, total body fat %, and FMI were like those observed for weight and BMI, with lower mean adiposity measurements in ART-conceived than NC offspring during childhood and adolescence, though with larger differences that were imprecisely estimated for several time points (**Figure 2**). As for weight and BMI, adiposity measures were higher in ART-conceived than NC adults, but with larger mean differences and wider CI’s that included the null (**Figure 2**).

**Figure 2.**
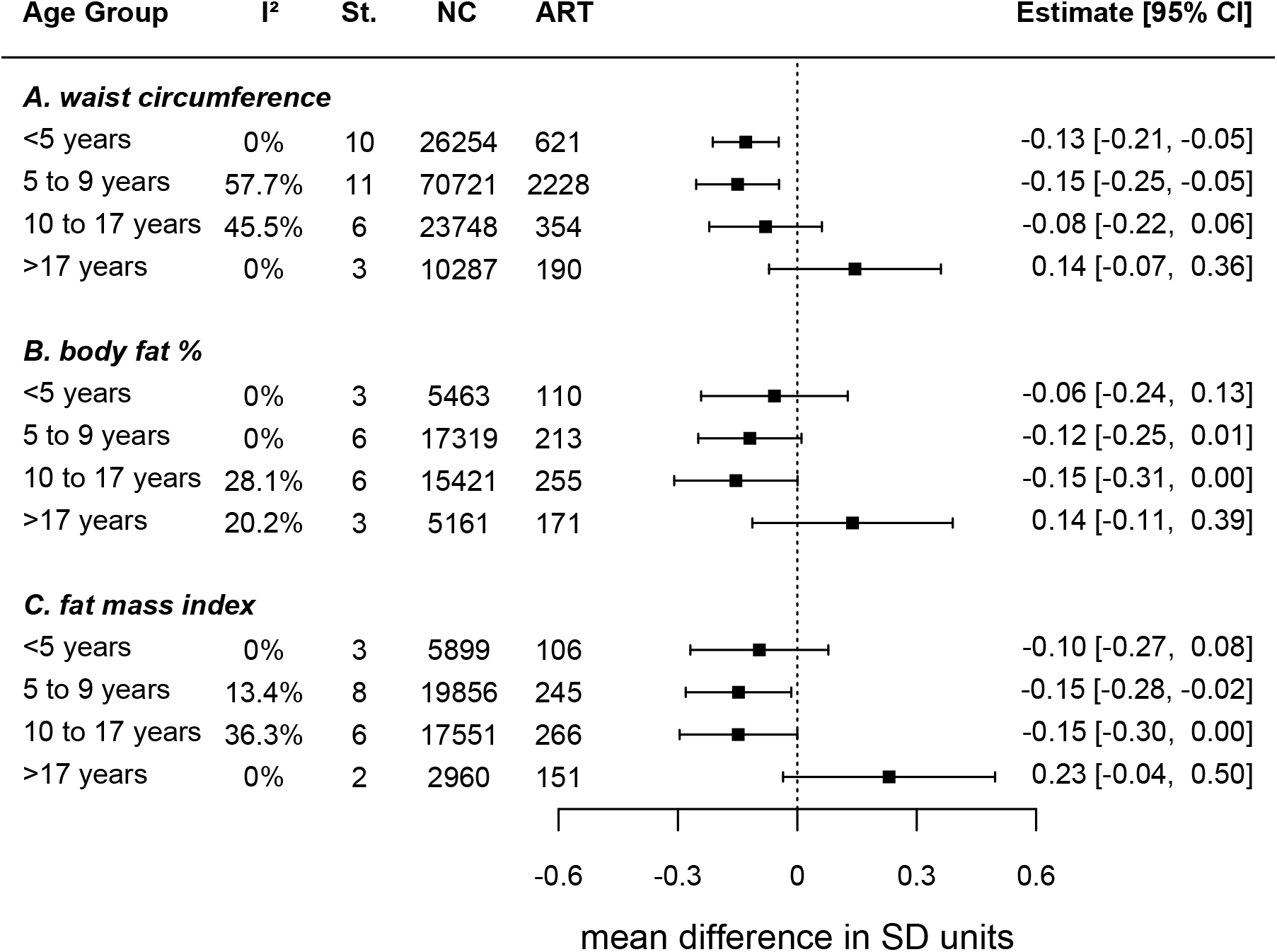
Parts A-C. Mean difference in (A) waist circumference, (B) body fat %, and (C) fat mass index between ART-conceived and NC offspring. Figure shows the pooled adjusted mean differences in SD units [and 95% confidence intervals] in (A) waist circumference, (B) body fat %, and (C) fat mass index at each age group between ART-conceived and NC offspring (ART minus NC). Cohort-specific results were adjusted for maternal age, parity, BMI, smoking, education, ethnicity/country of birth, plus offspring sex and age. St. is the number of cohort studies; NC is the number of NC ofspring; ART is the number of ART-conceived offspring; I² represents the percentage of total variability that is due to between cohort heterogeneity. Cohort-specific results are provided in eFigure 5-7.

Between-cohort heterogeneity was low to moderate for all outcomes at all ages, with a few exceptions. There was substantial between-cohort heterogeneity in results for length/height, weight, and BMI at ages <3 months and 3-5 months (**Figure 1**). Sensitivity analysis showed that results for outcomes at both ages were robust to influential cohorts, although they were slightly attenuated when the MoBa cohort was omitted (**eFigure 8**).

For some additional analyses there were too few ART conceptions to include all older age groups. Results were similar when ART was compared with sub-fertile NC and fertile NC (**Figure 3**), when ICSI and IVF were compared with NC (**Figure 4**), and in females and males (**eFigure 9**). Mean length/height, weight, and BMI were on average lower in those conceived by fresh ET compared with NC offspring across all available age groups i.e., from age <3 months to age 6 to 9 years (**Figure 5**). Conversely, differences were closer to the null for FET compared with NC, though results were imprecise (**Figure 5**). The differences in all growth and adiposity outcomes were only partially attenuated when analyses were restricted to singletons (**eFigure 10**), whereas differences between ART-conceived and NC offspring (**eFigure 11**), and between fresh ET and NC offspring (**eFigure 12**) were considerably attenuated after further adjustment for birth weight and gestational age, particularly at younger ages.

**Figure 3.**
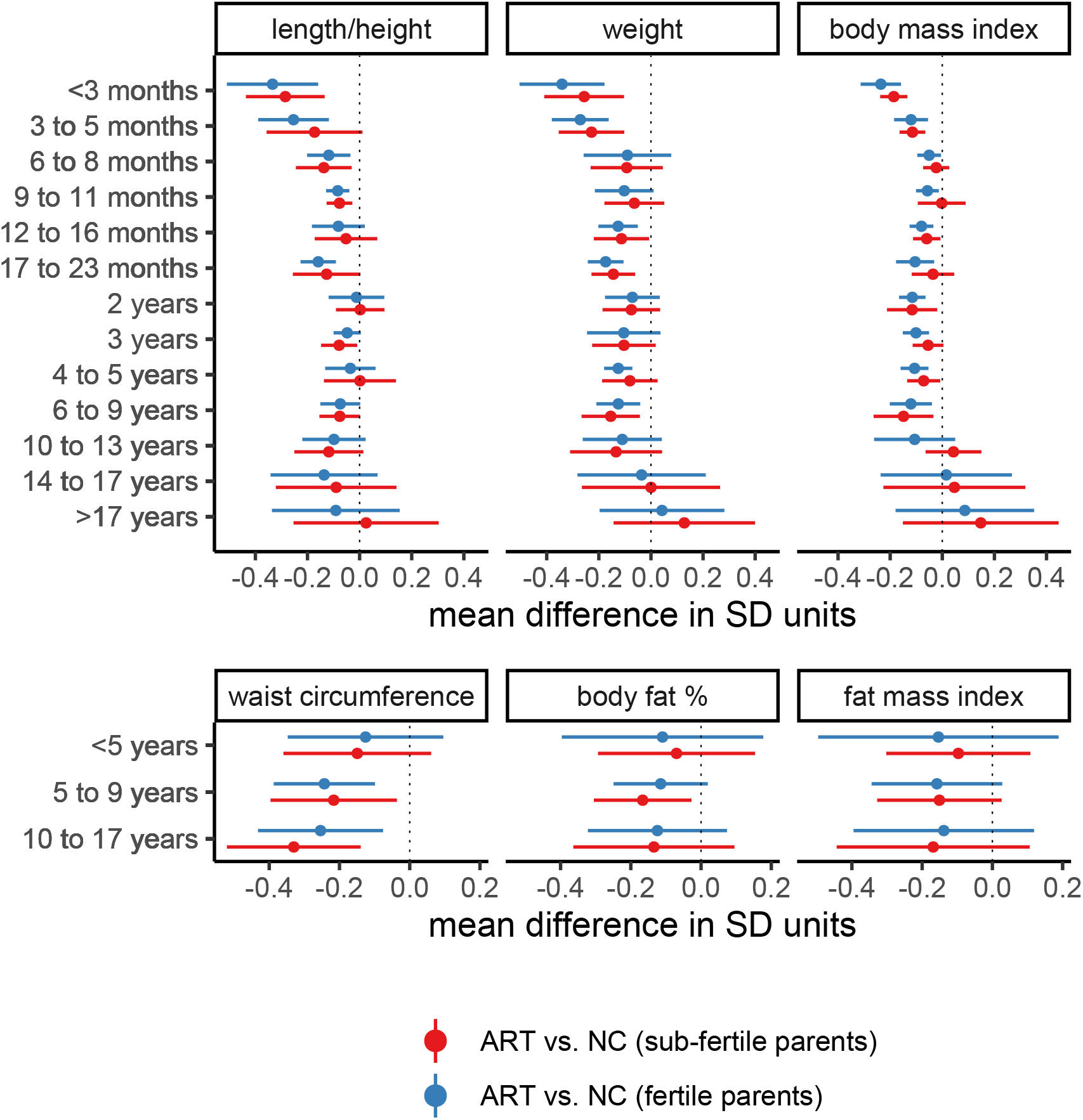
Mean difference in growth and adiposity outcomes between ART-conceived and NC offspring, separately for NC offspring of sub-fertile and fertile parents. Figure shows the pooled adjusted mean differences in SD units and 95% confidence intervals in growth and adiposity outcomes at each age group between ART-conceived and NC offspring (ART minus NC) of fertile parents (≤12 months to pregnancy and sub-fertile parents (>12 months to pregnancy). Cohort-specific results were adjusted for maternal age, parity, BMI, smoking, education, ethnicity/country of birth, plus offspring sex and age. The number of offspring at each age for the primary outcomes (length/height, weight and BMI) varied from 2,955 ART, 93,877 fertile NC, and 11,153 sub-fertile NC for weight at age 3 to 5 months to 51 ART, 3,350 fertile NC, and 494 sub-fertile NC for BMI at >17 years.

**Figure 4.**
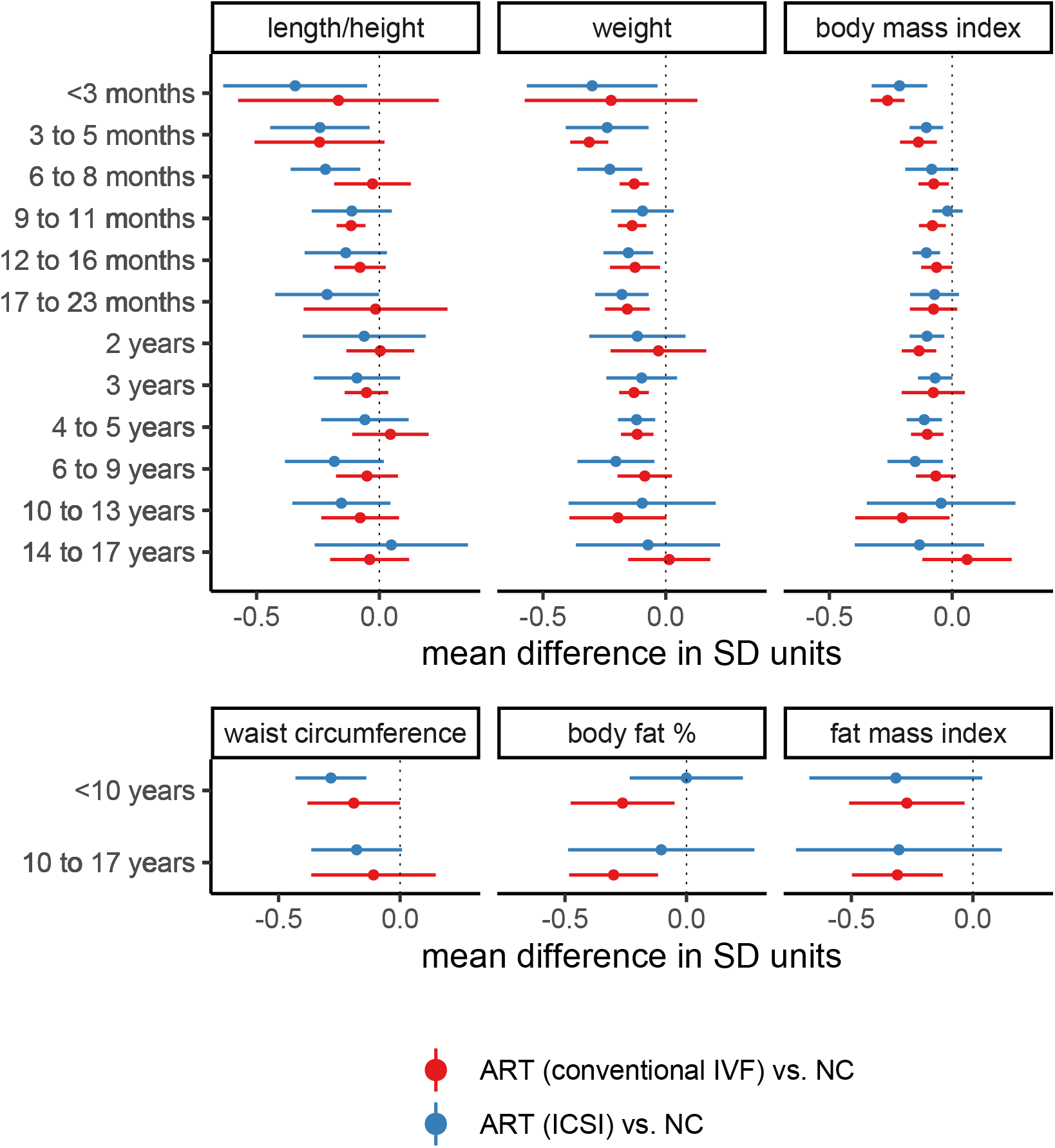
Mean difference in growth and adiposity outcomes between ART-conceived and NC offspring, separately for offspring conceived by conventional IVF and ICSI. Figure shows the pooled adjusted mean differences in SD units and 95% confidence intervals in growth and adiposity outcomes at each age group between ART-conceived and NC offspring (ART minus NC), separately for offspring conceived by conventional IVF and ICSI. Cohort-specific results were adjusted for maternal age, parity, BMI, smoking, education, ethnicity/country of birth, plus offspring sex and age. St. is the number of cohort studies. The number of offspring at each age for the primary outcomes (length/height, weight, and BMI) varied from 1,517 conventional IVF, 1,382 ICSI, and 102,386 NC for weight at age 3 to 5 months to 105 conventional IVF, 37 ICSI, and 11,164 NC for BMI at age 14 to 17 years.

**Figure 5.**
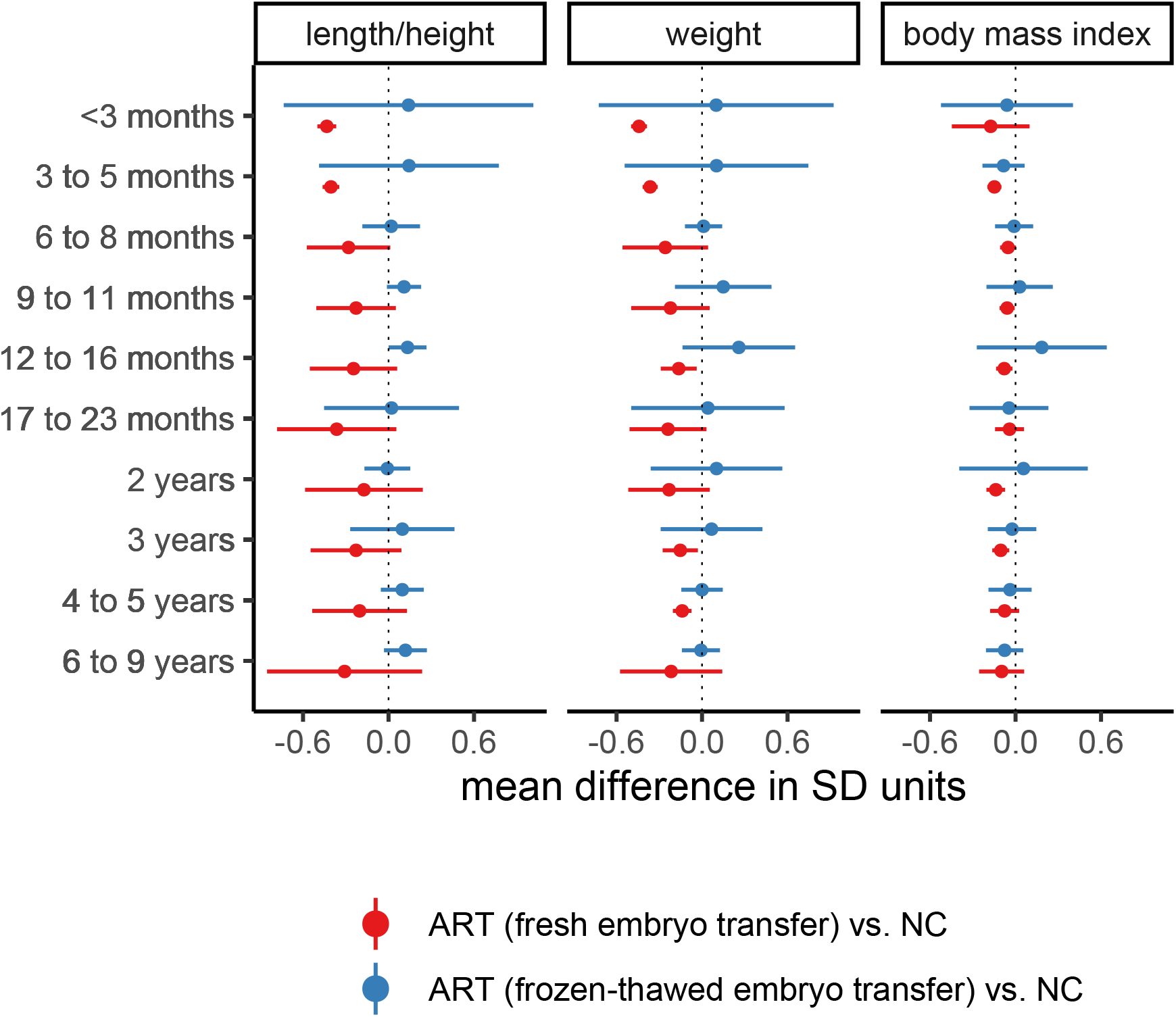
Mean difference in length/height, weight, and body mass index between ART-conceived and NC offspring, separately for offspring conceived using fresh and frozen-thawed embryo transfer. Figure shows the pooled adjusted mean differences in SD units and 95% confidence intervals in length/height, weight, and body mass index at each age group between ART-conceived and NC offspring (ART minus NC), separately for offspring conceived using fresh embryo transfer and frozen-thawed embryo transfer. Cohort-specific results were adjusted for maternal age, parity, BMI, smoking, education, ethnicity/country of birth, plus offspring sex and age The number of offspring at each age varied from 1,904 fresh embryo transfer, 303 frozen-thawed embryo transfer, and 78,128 NC for weight at age 3 to 5 months to 433 conventional fresh embryo transfer, 84 frozen-thawed embryo transfer, and 15,490 NC for BMI at age 17 to 23 months.

## DISCUSSION

We used data from 26 multinational cohort studies to investigate the association between ART conception and offspring growth and adiposity. The large number of offspring, and length of follow-up, allowed us to explore findings in subgroups by age from infancy to early adulthood. We found that offspring conceived by ART were on average shorter, lighter, and thinner from infancy to early adolescence than NC offspring. Differences were largest earlier in life but were small in magnitude across all ages. There was little evidence that differences were driven by parental subfertility given similar results when we compared ART with NC where parents conceived after 12 months and where conception occurred within a shorter period since start of trying. Those conceived from fresh embryos were smaller than NC offspring whereas frozen-thawed embryos were comparable to NC. Results appeared independent of multiple births and were at least partly mediated by birth weight and gestational age, particularly at younger ages.

Our findings are in line with previous studies and reviews of outcomes at birth and in young children (6, 15-17). Although not directly comparable with our study, our finding of smaller associations among older children is consistent with results from a recent study that found more rapid growth from birth to 3 years in ART-conceived than NC offspring (18). That study also examined outcomes at age 17 years in individuals screened for conscription in Norway and found no difference between ART-conceived and NC offspring at that age, which is consistent with our finding of no difference in growth in older adolescence.

Our results for fresh ET and FET are consistent with previous studies showing smaller birth weight in fresh ET compared with NC, and higher birth weight and large-for-gestational-age in FET compared with fresh ET (12, 13). Our study also agrees with results from a UK record linkage study that assessed birth size and body size at 6-8 weeks and 5 years in offspring born between 1997-2009, showing that compared with NC, offspring born by fresh embryos were lighter, and those born by FET were heavier at birth and 6-8 weeks, and that all groups had similar weight at 5 years (16).

The reasons for lower birth weight and higher risk of small-for-gestational-age shown in previous studies (8, 15) and the (modestly) smaller infant/child size in our study in ART-conceived offspring are not fully understood. The gametic and embryonic manipulations associated with ART may impact embryonic/fetal development in a manner that is reflected in different growth patterns relative to NC individuals. Growth differences could also reflect physiological responses to ART-induced lower birth size (and gestational age), and unlikely to be sex-specific or differ by ART type given our finding of broadly similar results in males and females and conventional IVF and ICSI. This is supported by our finding that differences attenuate by adjustment for birthweight and gestational age, though, this should be interpreted with caution since assumptions for such analyses and potential for collider bias makes them difficult to interpret (25, 26, 28). Other possible explanations include effects of ART-induced epigenetic alterations (29, 30), and effects of the ovarian stimulating hormones administered prior to ART. The different findings for fresh ET and FET may reflect effects of ovarian stimulation on endometrium and corpus luteum when using fresh embryos (31) or the impact of freezing on embryos.

Ours is the first study to date to explore long-term associations with waist circumference, body fat % and FMI, with results suggesting ART-conceived individuals had lower central and total adiposity in childhood, and possibly higher levels in adulthood. Our early life results agree with findings from 85 Singaporean ART-conceived offspring showing smaller skinfold thickness than NC offspring at 6 years (19). Our finding suggestive of an association between ART and higher adiposity in young adults agrees with results from a Nordic registry study showing slightly increased obesity risk in young ART-conceived adults (32). One possible reason for this result is that the rapid infant growth we observed in ART-conceived offspring continues (at a decelerating rate) for extended time. This is consistent with prior evidence of an association between rapid infant growth and adult overweight and obesity (33), and with cardiovascular disease risk in later adulthood (34).

It is important to note that our pooled effect sizes were small across all age groups, including at the youngest ages where they were largest in magnitude. For example, when expressed in its natural units, the largest differences in weight, observed at age <3 months, was 183 grams (95%CI: 105 to 261) lower in those conceived by ART. Therefore, it is unlikely that these differences will result in any clinically meaningful differences at any age. It is also worth acknowledging that our pooled results represent average differences in outcomes across all populations from all included cohorts, and there was some evidence of heterogeneity for some outcomes. However, sensitivity analyses indicated results were robust to influential cohorts and heterogeneity was due to differences in directionally consistent effect sizes.

Strengths of this study include the large sample size and inclusion of cohorts from different geographic regions which should make our findings generalisable to more populations. The large numbers allowed an assessment of heterogeneity in the main results, an exploration of potential roles of subfertility, different ART treatments, multiple births, and indirect effects through prematurity. Use of birth cohorts with comparison with NC children from the same underlying population as those conceived by ART and with identical baseline data collection, follow-up periods, and assessments in ART-conceived and NC children, is another important strength. Many previous studies compared clinical ART cohorts with a comparator group selected at the time of outcome assessment, thus lacking early data on potential confounders, and these were often selected from relatives/friends of the couples undergoing ART, which may introduce a selection bias (17). Record linkage studies mostly avoid this selection bias but are generally limited in the extent to which they can adjust for confounding or explore role of sub-fertility.

Limitations include low precision/power at older ages, which highlights the importance of measuring outcomes in adult life. Those with outcomes at older ages were exposed to ART some decades ago using treatments and embryo culture techniques that are less relevant to contemporary practices, thus making it difficult to know the extent to which findings would generalise to more recently born cohorts. Therefore, there is a need to promote collection of data on mode of conception from birth cohorts and to ensure those conceived by ART are included so that future analyses can continually add new cohorts to examine changes in associations by birth years and age. Our analysis was restricted to those with complete data on mode of conception, outcomes, and confounders which may have reduced precision of estimates and introduced bias due to missing data. Residual confounding by unmeasured factors (e.g., paternal health) is possible and might influence our findings.

In conclusion, we found that ART-conceived offspring were on average slightly smaller and had modestly lower adiposity than NC offspring during early life, with associations reduced with older child age, with some imprecise evidence for higher adiposity by early adulthood with ART conception. Overall, our findings are reassuring since differences in early growth were small though, there is a need for additional follow-up and studies with larger numbers into older ages to is investigate the possibility of greater adiposity in adulthood.

## Data Availability

All data produced in the present study are available upon reasonable request to the authors

## ACKNOWLEDGEMENTS

Cohort-specific acknowledgements and finding statements are provided in the supplement. The research leading to these results has received funding from the European Research Council under agreement No 101021566 (ART-HEALTH), the European Union’s Horizon 2020 research and innovation programme under grant agreement No 733206 (LifeCycle), the Medical Research Council (MC_UU_00011/6), British Heart Foundation (CH/F/20/90003 and AA/18/7/34219), and Bristol National Institute of Health Research Biomedical Research Centre (NF-0616-10102). DAL has received support from Medtronic and Roche Diagnostics for research unrelated to that presented here. SMN has received support from Roche Diagnostics, Access Fertility, Modern Fertility, Ferring Pharmaceuticals, TFP, and Merck for research unrelated to that presented here. All the other authors declare no competing interests. AE had full access to the data in the study and takes responsibility for the integrity of the data and the accuracy of the data analysis.

## REFERENCES

1. Zegers-Hochschild F, Adamson GD, Dyer S, et al. The International Glossary on Infertility and Fertility Care, 2017. Hum Reprod 2017; 32(9): 1786–801.

2. Crawford G, Ledger W. In vitro fertilisation/intracytoplasmic sperm injection beyond 2020. BJOG 2019; 126(2): 237–43.

3. Wang H, Abbas KM, Abbasifard M, et al. Global age-sex-specific fertility, mortality, healthy life expectancy (HALE), and population estimates in 204 countries and territories, 1950–2019: a comprehensive demographic analysis for the Global Burden of Disease Study 2019. Lancet 2020; 396(10258): 1160–203.

4. Levine H, Jørgensen N, Martino-Andrade A, et al. Temporal trends in sperm count: a systematic review and meta-regression analysis. Hum Reprod Update 2017; 23(6): 646–59.

5. Smith ADAC, Tilling K, Nelson SM, Lawlor DA. Live-birth rate associated with repeat in vitro fertilization treatment cycles. JAMA 2015; 314(24): 2654–62.

6. Goisis A, Remes H, Martikainen P, Klemetti R, Myrskylä M. Medically assisted reproduction and birth outcomes: a within-family analysis using Finnish population registers. Lancet 2019; 393(10177): 1225–32.

7. Pinborg A, Wennerholm UB, Romundstad LB, Loft A, Aittomaki K, Söderström-Anttila V, et al. Why do singletons conceived after assisted reproduction technology have adverse perinatal outcome? Systematic review and meta-analysis. Hum Reprod Update. 2013;19(2):87–104.

8. Berntsen S, Söderström-Anttila V, Wennerholm U-B, Laivuori H, Loft A, Oldereid NB, et al. The health of children conceived by ART: ‘the chicken or the egg?’. Hum Reprod Update. 2019;25(2):137–58.

9. Dhalwani NN, Boulet SL, Kissin DM, Zhang Y, McKane P, Bailey MA, et al. Assisted reproductive technology and perinatal outcomes: conventional versus discordant-sibling design. Fertil Steril. 2016;106(3):710–6.e2.

10. Esteves SC, Roque M, Bedoschi G, Haahr T, Humaidan P. Intracytoplasmic sperm injection for male infertility and consequences for offspring. Nat Rev Urol. 2018;15(9):535–62.

11. Smith A, Tilling K, Lawlor DA, Nelson SM. Live birth rates and perinatal outcomes when all embryos are frozen compared with conventional fresh and frozen embryo transfer: a cohort study of 337,148 in vitro fertilisation cycles. BMC Med. 2019;17(1):202.

12. Terho AM, Pelkonen S, Opdahl S, Romundstad LB, Bergh C, Wennerholm UB, et al. High birth weight and large-for-gestational-age in singletons born after frozen compared to fresh embryo transfer, by gestational week: a Nordic register study from the CoNARTaS group. Hum Reprod. 2021;36(4):1083–92.

13. Westvik-Johari K, Romundstad LB, Lawlor DA, Bergh C, Gissler M, Henningsen A-KA, et al. Separating parental and treatment contributions to perinatal health after fresh and frozen embryo transfer in assisted reproduction: A cohort study with within-sibship analysis. PLOS Med. 2021;18(6):e1003683.

14. Shi Y, Sun Y, Hao C, Zhang H, Wei D, Zhang Y, et al. Transfer of Fresh versus Frozen Embryos in Ovulatory Women. N Engl J Med. 2018;378(2):126–36.

15. Hart R, Norman RJ. The longer-term health outcomes for children born as a result of IVF treatment: Part I--General health outcomes. Hum Reprod Update. 2013;19(3):232–43.

16. Hann M, Roberts SA, D’Souza SW, Clayton P, Macklon N, Brison DR. The growth of assisted reproductive treatment-conceived children from birth to 5 years: a national cohort study. BMC Med. 2018;16(1):224.

17. Guo XY, Liu XM, Jin L, Wang TT, Ullah K, Sheng JZ, et al. Cardiovascular and metabolic profiles of offspring conceived by assisted reproductive technologies: a systematic review and meta-analysis. Fertil Steril. 2017;107(3):622–31.e5.

18. Magnus MC, Wilcox AJ, Fadum EA, Gjessing HK, Opdahl S, Juliusson PB, et al. Growth in children conceived by ART. Hum Reprod. 2021;36(4):1074–82.

19. Huang JY, Cai S, Huang Z, Tint MT, Yuan WL, Aris IM, et al. Analyses of child cardiometabolic phenotype following assisted reproductive technologies using a pragmatic trial emulation approach. Nat Commun. 2021;12(1):5613.

20. von Elm E, Altman DG, Egger M, Pocock SJ, Gotzsche PC, Vandenbroucke JP. The Strengthening the Reporting of Observational Studies in Epidemiology (STROBE) statement: guidelines for reporting observational studies. Lancet. 2007;370(9596):1453–7.

21. Jaddoe VWV, Felix JF, Andersen AN, Charles MA, Chatzi L, Corpeleijn E, et al. The LifeCycle Project-EU Child Cohort Network: a federated analysis infrastructure and harmonized data of more than 250,000 children and parents. Eur J Epidemiol. 2020;35(7):709–24.

22. Pinot de Moira A, Haakma S, Strandberg-Larsen K, van Enckevort E, Kooijman M, Cadman T, et al. The EU Child Cohort Network’s core data: establishing a set of findable, accessible, interoperable and re-usable (FAIR) variables. Eur J Epidemiol. 2021;36(5):565–580.

23. Voerman E, Santos S, Inskip H, Amiano P, Barros H, Charles MA, et al. Association of gestational weight gain with adverse maternal and infant outcomes. JAMA. 2019;321(17):1702–15.

24. Carson SA, Kallen AN. Diagnosis and management of infertility: a review. JAMA. 2021;326(1):65–76.

25. Pearce N, Lawlor DA. Causal inference—so much more than statistics. Int J Epidemiol. 2017;45(6):1895–903.

26. Lipsky AM, Greenland S. Causal Directed Acyclic Graphs. JAMA. 2022;doi:10.1001/jama.2022.1816

27. Higgins JP, Thompson SG, Deeks JJ, Altman DG. Measuring inconsistency in meta-analyses. Bmj. 2003;327(7414):557–60.

28. Munafo MR, Tilling K, Taylor AE, Evans DM, Davey Smith G. Collider scope: when selection bias can substantially influence observed associations. Int J Epidemiol. 2018;47(1):226–35.

29. Novakovic B, Lewis S, Halliday J, Kennedy J, Burgner DP, Czajko A, et al. Assisted reproductive technologies are associated with limited epigenetic variation at birth that largely resolves by adulthood. Nat Commun. 2019;10(1):3922.

30. Caramaschi D, Jungius J, Page CM, Novakovic B, Saffery R, Halliday J, et al. Association of medically assisted reproduction with offspring cord blood DNA methylation across cohorts. Hum Reprod. 2021; 36(8):2403–2413.

31. Shapiro BS, Daneshmand ST, Garner FC, Aguirre M, Hudson C, Thomas S. Evidence of impaired endometrial receptivity after ovarian stimulation for in vitro fertilization: a prospective randomized trial comparing fresh and frozen–thawed embryo transfer in normal responders. Fertil Steril. 2011;96(2):344–8.

32. Norrman E, Petzold M, Gissler M, Spangmose AL, Opdahl S, Henningsen A-K, et al. Cardiovascular disease, obesity, and type 2 diabetes in children born after assisted reproductive technology: A population-based cohort study. PLOS Med. 2021;18(9):e1003723.

33. Monteiro POA, Victora CG. Rapid growth in infancy and childhood and obesity in later life – a systematic review. Obes Rev. 2005;6(2):143–54.

34. Eriksson JG, Forsen T, Tuomilehto J, Winter PD, Osmond C, Barker DJ. Catch-up growth in childhood and death from coronary heart disease: longitudinal study. BMJ. 1999;318(7181):427–31.

