## Supplementary material for "Association of assisted reproductive technology with offspring growth and adiposity from infancy to early adulthood"

[eFigure 8. Mean difference in length/height, weight, and body mass index between ART-conceived and NC offspring at ages <3 months and 3-5 months, after leaving each cohort study out of the meta-analysis (to identify influential cohorts)	33](#_Toc98667454)

### **eMethods**

This section provides a description of the 26 cohort studies, including details of how exposures/outcomes were measured and how ethical approvals and consent were obtained. eTable 1 summarises the characteristics of the cohort studies. eTable 2 provides descriptive data on the numbers included in the analysis including for each timepoint where repeats are available and for each exposure and outcome, as well as the means and standard deviations of each outcome and age at outcome assessment at each assessment wave.

**1. All Our Families Study (AOF)**

AOF is a community-based longitudinal pregnancy cohort of mother-child dyads investigating maternal, birth and child development outcomes^1^. A total of 3387 pregnant women, residing in Calgary, Canada, enrolled in the study between 2008-2011. Data are collected through self-report questionnaires during pregnancy, post-partum, and post-birth. Mothers have completed eight questionnaires to date spanning pregnancy to 8 years post-birth and provided access to their labour and delivery medical records.

Up to 41 offspring conceived by ART and 1,780 NC offspring were included in this study (including multiple births). AOF contributed results to the main analysis (ART vs. NC) and to additional analysis stratified by sex, sub-fertility, IVF/ICSI, and ET/FET (for FET only), for height, weight, and BMI. Data were available for all study confounders (i.e., maternal age, parity, BMI, smoking, education, ethnicity and offspring sex and age at outcome assessment).

**2. Amsterdam Born Children and their Development Study (ABCD)**

Between January 2003 and March 2004, all pregnant women living in Amsterdam were asked to participate in the ABCD study during their first prenatal visit to an obstetric care provider (general practitioner, midwife, or gynaecologist)^2^. Of the 12,373 women approached, 8,266 women filled out the pregnancy questionnaire (response rate: 67%). Of this group, 7,050 women granted permission for follow-up (85%) and 7,043 women granted permission for perusal of her and her child’s medical files (85%). Through a questionnaire, women provided information on time to pregnancy (in months) and mode of conception.

Up to 61 ART-conceived offspring and 4,701 NC offspring were included in this study (singleton births only). ABCD contributed results to the main analysis (ART vs. NC) and to additional analysis stratified by sex, sub-fertility, and IVF/ICSI, for all study outcomes (i.e., height, weight, BMI, waist circumference, body fat % and fat mass index). Data were available for all study confounders (i.e., maternal age, parity, BMI, smoking, education, ethnicity and offspring sex and age at outcome assessment). Confounders were measured by questionnaire (maternal self-report) administered during first trimester of pregnancy.

**3. Avon Longitudinal Study of Parents and Children (ALSPAC)**

ALSPAC is a prospective birth cohort study that recruited all pregnant women residing within the catchment area of 3 National Health Service authorities in southwest England with an expected date of delivery between April 1991 and December 1992^3-5^. The initial number of pregnancies enrolled is 14,541 (for these at least one questionnaire has been returned or a “Children in Focus” clinic had been attended by 19/07/99). Of these initial pregnancies, there was a total of 14,676 fetuses, resulting in 14,062 live births and 13,988 children who were alive at 1 year of age. Please note that the study website contains details of all the data that is available through a fully searchable data dictionary and variable search tool" and reference the following webpage:

<http://www.bristol.ac.uk/alspac/researchers/our-data/>.

Detailed information has been collected from offspring and their parents using questionnaires, data extraction from medical records, linkage to health records, and clinic assessments up to the last completed contact. Numerous height and weight measures have been obtained from various sources from after birth to age 25 years, including from routine data collected from midwives, health visitors, linkage to child health records, and from ALSPAC research clinic visits. Waist circumference was measured to the nearest mm at research clinics. Body fat % was measured by bio-electrical-impedance (Tanita Body Fat Analyser), and fat mass was measured by whole-body DXA scans (Lunar prodigy).

Up to 56 offspring conceived by ART and 10,380 NC offspring were included in this study (including multiple births). ALSPAC contributed results to the main analysis (ART vs. NC) and to additional analyses stratified by sex, and sub-fertility, for all study outcomes (i.e., height, weight, BMI, waist circumference, body fat % and fat mass index). Data were available for all study confounders (i.e., maternal age, parity, BMI, smoking, education, ethnicity and offspring sex and age at outcome assessment).

**4. Babies After SCOPE: Evaluating the Longitudinal Impact on Neurological and Nutritional Endpoints (BASELINE)**

The Cork BASELINE Birth Cohort Study ^6^ is the first Irish prospective birth cohort study and provides detailed information on maternal health, fetal growth, childhood nutrition, growth, and development in the first five years of life. Participants were healthy nulliparous women with singleton pregnancies recruited from the Screening for Pregnancy Endpoints (SCOPE) pregnancy cohort. Detailed information, including information on demographics, lifestyle, obstetric history (including history of fertility and ART) and maternal anthropometric assessment were collected by research midwives at 15 weeks and 20 weeks’ gestation using questionnaires and clinical examination. Child anthropometric measures were completed at each study visit (at day 2 and at age 2, 6 and 12 months, and 2 and 5 years) and were conducted according to standard operating procedures. Body composition was measured using air displacement plethysmography with the PEA POD Infant Body Composition System (COSMED USA, Concord, CA).

Up to 20 ART-conceived offspring and 1,031 NC offspring were included in this study (singleton births only). BASELINE contributed results to the main analysis (ART vs. NC) and to additional analysis stratified by sex and sub-fertility, for all study outcomes (i.e., height, weight, BMI, waist circumference, body fat % and fat mass index). Data were available for all study confounders (maternal age, BMI, smoking, education, ethnicity and offspring sex and age at outcome assessment), except for parity as only nulliparous women were included.

**5. Barwon Infant Study (BIS)**

BIS is a prospective pre-birth population-based cohort study (n = 1064 mother–1074 infant pairs [10 sets of twins]) with antenatal recruitment conducted in the Barwon region in Victoria, Australia^7^. Pregnant women were recruited before 28 weeks of gestation between years 2010 and 2013. Detailed questionnaire and clinical data and extensive biospecimens have been collected from multiple time points from pregnancy to 4 years of age. BMI was calculated from height and weight measured by research staff during clinic visits. Body fat percentage was measured using DXA scanning at 4 years of age.

Up to 35 offspring conceived by ART and 673 NC offspring were included in this study. BIS contributed results to the main analysis (ART vs. NC) and to additional sex-stratified analysis, for height, weight, BMI, and body fat % at the 12-month and 4-year time points. Data were available for all study confounders (i.e., maternal age, parity, BMI, smoking, education, ethnicity and offspring sex and age at outcome assessment).

**6. Born in Guangzhou Cohort Study (BIGCS)**

BIGCS a prospective birth cohort study launched by the Guangzhou Women and Children’s Medical Center (GWCMC), China, in 2012^8^. Pregnant women attending their first antenatal care visit at the GWCMC were invited to participate in BIGCS if they were at <20 weeks of gestation, if they intended to deliver at GWCMC and if they intended to stay in Guangzhou for at least 3 years after delivery. Data on the exposures (ART) were collected using a self-reported questionnaire at recruitment. Weight and length/height were measured by trained research assistants. The children were asked to remove their shoes and heavy clothes but to retain a single layer of clothing.

Up to 379 ART-conceived offspring and 9,875 NC offspring were included in this analysis (singleton births only). BIGCS contributed results to the main analysis (ART vs. NC) and to additional analysis stratified by sex and sub-fertility, for, height, weight, and BMI. Data were available for all study confounders (maternal age, BMI, smoking, education, parity and offspring sex and age at outcome assessment), except for ethnicity, which was not inlcuded in the model as all participants were Chinese and about 98% were Han Chinese. Data on potential confounders were collected using self-reported questionnaires.

**7. Clinical review of the Health of 22–33 years old conceived with and without ART (CHART)**

The CHART study^9^ is a clinical review of a cohort comprising 547 ART-conceived adults and 549 matched naturally conceived (NC) controls. Recruitment was by letter (postal mailing), with a follow-up letter after 3 weeks and a phone call after a further 3 weeks. Additional attempts at contacting the participants included the use of social media and phone calls to their mothers. Data on clinical and biomarker outcomes were measured including cardiovascular structure and function, auxology, respiratory function, cardiometabolic profile and epigenome-wide DNA methylation analysis. Confounding variables were collected via a questionnaire.

Up to 130 ART-conceived and 73 NC offspring were included in this study. CHART contributed results to the main analysis (ART vs. NC) and to additional analysis stratified by sex and ET/FET, for all study outcomes (i.e., height, weight, BMI, waist circumference, body fat % and fat mass index). Data were available for some study confounders (maternal age, parity, education, and offspring sex and age at outcome assessment), but not for maternal BMI, smoking or ethnicity.

**8. Danish National Birth Cohort (DNBC)**

DNBC is a nationwide cohort of pregnant women, recruited from 1996 through 2002 consisting of 100,415 pregnancies^10^. Information on lifestyle and environmental factors potentially associated with offspring health was collected through 4 prenatal and postnatal telephone interviews at target ages gestational weeks 12 and 30 and child ages 6 and 18 months. The parent-child dyads were then invited for follow-up at 7, 11, and 18 years.

Up to 1481 offspring conceived by ART and 45,203 NC offspring were included in this study (including multiple births). DNBC contributed results to the main analysis (ART vs. NC) and to additional sex-stratified analysis, for height, weight, BMI, and waist circumference. Data were available for most study confounders (i.e., maternal age, parity, BMI, smoking, education, and offspring sex and age at outcome assessment), except ethnicity (though >95% were of white ethnicity).

**9. Etude de cohorte généraliste, menée en France sur les Déterminants pré et post natals précoces du développement psychomoteur et de la santé de l’Enfant (EDEN)**

EDEN is a birth cohort study that enrolled 2,002 pregnant women attending their prenatal visit before 24 weeks' gestation at Nancy and Poitiers university hospitals (France) between 2003 and 2006^11^. Detailed information has been collected from parents using questionnaires (including mode of conception and fertility treatment), data extraction from obstetrical file and three clinic assessments up to the last completed contact at age 11 yrs. Height and weight measures were obtained from birth to age 9 years mainly through parental report of measurements performed by health professionals retrieved from the child health booklet and from research clinic visits at 1,3 and 5 years. Waist circumference was measured at the 3- and 5-year research clinic visits. Body fat % was measured by bio-impedance at 5 years.

Up to 22 ART-conceived offspring and 1,326 NC offspring were included in this study. EDEN contributed results to the main analysis (ART vs. NC) and to additional analysis stratified by sex and sub-fertility, for all study outcomes (i.e., height, weight, BMI, waist circumference, body fat % and fat mass index). Data were available for all study confounders (maternal age, BMI, smoking, education, ethnicity, parity and offspring sex and age at outcome assessment).

**10. Etude Longitudinale Françcaise depuis l’Enfance (ELFE)**

ELFE is a nationwide birth cohort, including 18 329 children born in 2011 in a random sample of 349 maternity units from mainland France^12^. Inclusion criteria were singleton or twins born after 33 weeks' gestation to mothers aged ≥18 years and not planning to move outside of metropolitan France in the next 3 years. Detailed information has been collected from parents using questionnaires ((including mode of conception and fertility treatment), data extraction from obstetrical file, and clinical assessment by general practitioner at age 2. Height and weight measures have been collected through parental report of measurements performed by health professionals retrieved from the child health booklet and from the 2-yr general practionner clinical exam. Data up to 39 months have been used for this analysis.

Up to 309 ART-conceived offspring and 9,632 NC offspring were included in this study (including multiple births). ELFE contributed results to the main analysis (ART vs. NC) and to additional analysis stratified by sex, ICSI/IVF, and sub-fertility, for height, weight, and BMI. Data were available for all study confounders (maternal age, BMI, smoking, education, ethnicity, parity and offspring sex and age at outcome assessment).

**11. EU Childhood Obesity Project (CHOP)**

The European Childhood Obesity Project (CHOP) was a one-year multicentre double-blind randomized controlled intervention trial including 1678 children (registered at ClinicalTrials.gov: NCT00338689). Healthy singleton term infants born between 1^st^ October 2002 and 31^st^ July 2004 were recruited in five European countries (Belgium, Germany, Italy, Poland, Spain) during their first 8 weeks of life. They were randomized to cow-milk based formula with either higher or lower protein-content. Additionally, a reference group of breastfed children was included. The aim was to test whether feeding infant formula, which differ in their level of milk proteins, can influence infant growth and the risk of later childhood obesity ('early protein hypothesis'). After the intervention, children were prospectively followed up until the age of 11 years. More detailed information on the study design and results can be found elsewhere ^13-16^. Data on the exposure (ART) and the confounders were collected using questionnaires at baseline. Study nurses measured height, weight and waist circumference of the children at various ages.

Up to 20 ART-conceived offspring and 1,479 NC offspring were included in this study (singleton births only). CHOP contributed results to the main analysis (ART vs. NC) only, for height, weight, BMI, and waist circumference. Data were available for all study confounders (maternal age, BMI, smoking, education, ethnicity, parity and offspring sex and age at outcome assessment).

**12. Gene and Environment: Prospective Study on Infancy in Italy (GASPII)**

GASPII is a prospective birth cohort study of 709 children born between June 2003 and October 2004 in 2 maternal units located in Rome, Italy^17^.

Up to 8 ART-conceived offspring and 554 NC offspring were included in this study. GASPII contributed results to the main analysis (ART vs. NC) to additional analysis stratified by sub-fertility, for height, weight, BMI, and waist circumference. Data were available for all study confounders (maternal age, BMI, smoking, education, ethnicity, parity and offspring sex and age at outcome assessment).

**13. Generation R (Gen-R)**

Gen-R is a population-based prospective cohort study from fetal life until adulthood^18,19^. In total, 9,778 mothers with a delivery date from April 2002 until January 2006 were enrolled in the study. Response at baseline was 61%. Extensive assessments including physical examinations and DXA measurements are performed in mothers, fathers and their children.

Up to 47 ART-conceived offspring and 4,334 NC offspring were included in this study. Gen-R contributed results to the main analysis (ART vs. NC) to additional analysis stratified by sex, for height, weight, BMI, and fat mass index. Data were available for all study confounders (maternal age, BMI, smoking, education, ethnicity, parity and offspring sex and age at outcome assessment).

**14. Generation XXI (G21)**

Generation XXI (G21) is a prospective population-based birth cohort that recruited pregnant women delivering live-born infants (including multiple births) between April 2005 and August 2006 at all five public maternity units that served the metropolitan area of Porto, Portugal^20^. Overall, 8,647 infants with gestational age above 23 weeks and their mothers (n=8,495) were enrolled (91.4% participation). Subsequent evaluations of the entire cohort took place when children were 4 (n=7,459), 7 (n=6,889), 10 (n=6,397), and 13 years old (n=4,640, interrupted due to the COVID-19 pandemic). The cohort has more than 95% Caucasian participants. Data on demographic and socioeconomic characteristics, lifestyles, obstetric history, and anthropometrics were collected within 72 hours after delivery, in a face-to-face interview conducted by trained interviewers using structured questionnaires. During follow-up, physical examination and multiple questionnaires were performed by trained examiners, according to standard procedures, including weight, height and waist circumference measurements, body fat % and fat mass was measured by bio-impedance.

Up to 92 ART-conceived offspring and 5,746 NC offspring were included in this study (including multiple births). G21 contributed results to the main analysis (ART vs. NC) to additional analysis stratified by sex, sub-fertility, and IVF/ICSI, for all study outcomes (i.e., height, weight, BMI, waist circumference, body fat % and fat mass index). Data were available for all study confounders (maternal age, BMI, smoking, education, maternal country of birth, parity and offspring sex and age at outcome assessment).

**15. Growing Up in Ireland Infant Cohort (GUI)**

GUI The Growing Up in Ireland (GUI) study is a nationally representative prospective infant cohort study which recruited a random sample of 11,134 infants born in Ireland from 2007-2008^21^. The children and their families had a baseline face-to-face questionnaire-based interview conducted by trained interviewers in participating households when the infants were approximately nine months old. Mother-infant pairs were subsequently followed-up by home interview when infants were three and five years old. The child’s height and weight were measured by a trained interviewer using a validated standard measuring stick (Leicester portable height measure) and a medically approved weighing scale (SECA 835 digital weighing scales). Parity defined as the total number of stillbirths and live births a woman has had was not available, however, we used the number of individuals currently in the study household who were a son/daughter of the mother as a proxy for parity.

Up to 173 ART-conceived offspring and 9,742 NC offspring were included in this study (including multiple births). GUI contributed results to the main analysis (ART vs. NC) to additional analysis stratified by sex, and IVF/ICSI, for height, weight, and BMI. Data were available for all study confounders (maternal age, BMI, smoking, education, ethnicity, parity and offspring sex and age at outcome assessment).

**16. Growing Up in New Zealand (GUiNZ)**

GUiNZ is a prospective birth cohort study that recruited 6,853 children via their pregnant mothers if they had an expected delivery date between 25 April 2009 and 25 March 2010 and were residing within a geographically defined region of New Zealand which was chosen because it could provide a cohort of births that would be representative of all current births in NZ, especially with respect to ethnic and socioeconomic diversity^22^. Birth parameters were retrieved via linkage to routine perinatal records (with maternal consent) and repeated child height and weight measurements were collected as part of field interviews when the children were 2 years and 4 years of age. Anthropometric measurements were undertaken by trained interviewers using a standardised approach used by the NZ Ministry of Health.

Up to 173 ART-conceived offspring and 4,274 NC offspring were included in this study. GUiNZ contributed results to the main analysis (ART vs. NC) to additional analysis stratified by sex, for height, weight, BMI, and waist circumference. Data were available for all study confounders (maternal age, BMI, smoking, education, ethnicity, parity and offspring sex and age at outcome assessment).

**17. Growing up in Singapore Towards healthy Outcomes (GUSTO)**

GUSTO recruited pregnant women aged 18 years and above, attending their first trimester antenatal dating ultrasound scan clinic at Singapore’s two major public maternity units^23^. Women were eligible if 18 years and older, Singaporean citizens or permanent residents, with self-reported homogenous ethnic ancestry (Chinese, Indian, Malay), intended to deliver at the either of the recruitment hospitals and reside in Singapore for the next 5 years. Women greater than 14 weeks of gestation, receiving chemotherapy, psychotropic medications, or having an existing type I diabetes mellitus diagnosis at the time of recruitment were excluded. Women who ultimately did not agree to donate birth tissues (cord, placenta, cord blood) were also excluded. Women were asked to self-report whether the current pregnancy was conceived via IVF and use of assisted reproductive technologies, along with relevant treatment modalities, were confirmed via medical record review by a senior obstetrician and fertility consultant. Women reporting IVF conception with multiple gestations were further excluded.

Maternal obstetric and medical history including self-reported pre-pregnancy body weight, sociodemographic characteristics, and health behaviors, such as personal and family tobacco smoking, were ascertained by study staff administered standardized questionnaire at recruitment and at a study visit at 26-28 weeks gestation. Mode of delivery, procedures, and complications and birth weight, length, and head circumference were abstracted from delivery record. At all post-delivery visits weight (calibrated Seca 334 or Seca 803 digital scales; Seca, Hamburg, Germany); recumbent crown-to-sole length (up to 24 months; Seca 210 Mobile Measuring Mat) / standing height (beginning at 18 months; Seca 213 Stadiometer); head, mid-upper arm, and abdominal circumferences (inelastic measuring tape); and skinfold (triceps, biceps, subscapular, and suprailiac) thickness (Holtain skinfold calipers; Holtain Ltd., Crymych, UK) were collected by trained study staff in duplicate or triplicate (or 4-5 time for skinfold) and averaged under standardized protocols based on U.S. National Health and Nutrition Examination Survey (NHANES) protocols.

Up to 66 ART-conceived offspring and 935 NC offspring were included in this study (singletons only). GUSTO contributed results to the main analysis (ART vs. NC) to additional analysis stratified by sex, IVF/ICSI, and ET/FET for all study outcomes (i.e., height, weight, BMI, waist circumference, body fat % and fat mass index). Data were available for all study confounders (maternal age, BMI, smoking, education, ethnicity, parity and offspring sex and age at outcome assessment).

**18. Healthy Growth Study (HGS)**

HGS is a child cohort study started in 2007 that recruited schoolchildren aged 9–13 years, attending primary schools located in municipalities within the counties of Attica, Aitoloakarnania, Thessaloniki and Iraklio, in Greece^24^. Participants underwent a physical examination by two trained members of the research team. The protocol and equipment used were the same in all schools.

Weight was measured to the nearest 10 g using a digital scale (Seca Alpha, model 770; Seca, Hamburg, Germany). Children were weighed without shoes in the minimum clothing possible. Height was measured to the nearest 0·1 cm using a commercial stadiometer (Leicester Height Measure; Invicta Plastics, Oadby, UK) with the child standing barefoot, keeping shoulders in a relaxed position, arms hanging freely and head in the Frankfurt horizontal plane. Waist circumference was measured to the nearest 0·1 cm with the use of a non-elastic tape (Hoechstmass, Sulzbach, Germany) with the child standing, at the end of a gentle expiration, after placing the measuring tape on a horizontal plane around the trunk, at the level of the umbilicus, midway between the lower rib margin and the iliac crest. Bioelectrical impedance analysis (BIA) was used for the assessment of percentage body fat (Akkern BIA 101; Akkern Srl, Florence, Italy). Data on the socio-economic background of the families having at least one child participating in the study were collected from the parents (most preferably from the mother) during scheduled face-to-face interviews at school.

Up to 63 ART-conceived offspring and 2,182 NC offspring were included in this study. HGS contributed results to the main analysis (ART vs. NC) to additional analysis stratified by sex, and IVF/ICSI, for all study outcomes (i.e., height, weight, BMI, waist circumference, body fat % and fat mass index). Data were available for all study confounders (maternal age, BMI, smoking, education, ethnicity, parity and offspring sex and age at outcome assessment).

**19. Italian Twins Register (ITR)**

ITR^25^ is a population-based registry of voluntary twins. Since its inception, 29,000 twins have been enrolled and about 20% of them are minors. The ITR collects information on a large spectrum of phenotypes by either self-reported questionnaires or clinical examinations. In the case of underage twins, the information is reported by the parents who sign an informed consent form.

The ITR offspring were aged between 6 months and 13 years at time of outcome assessments. Due to the wide age range at outcome assessment, ITR was analysed in 7 separate age groups that each included between 32 and 54 ART-conceived offspring (and between 140 and 819 NC offspring (multiple births only). ITR contributed results to the main analysis (ART vs. NC) to additional analysis stratified by sex, and IVF/ICSI (for IVF only), for height, weight, and BMI. Data were available for all study confounders (maternal age, BMI, smoking, education, parity and offspring sex and age at outcome assessment) except for ethnicity.

**20. Millenium Cohort Study (MCS)**

MCS is a nationally representative birth cohort study that followed 19,244 children born in the UK in 2000–2002^26^. Baseline interviews were conducted when the children were approximately nine months old, and follow-up interviews were conducted when the children were around 3, 5, 7, 11, 14 and 17 years old. MCS includes detailed information about the demographic, health and socio-economic characteristics of the respondents and their families. At the baseline interview, the cohort member’s mother was asked whether they had used any fertility treatment to conceive.

Up to 30 ART-conceived offspring and 2,153 NC offspring were included in this study (including multiple births). MCS contributed results to the main analysis (ART vs. NC) to additional analysis stratified by sex, sub-fertility, and IVF/ICSI, for all study outcomes (i.e., height, weight, BMI, waist circumference, body fat % and fat mass index). Data were available for all study confounders (maternal age, BMI, smoking, education, ethnicity, parity and offspring sex and age at outcome assessment).

**21. MUltiple BIrth Cohort Study (MUBICOS)**

MUBICOS ^25^ has been established within the Italian Twin registry since 2010 but these cohorts do not overlap. About 360 families were enrolled and DNA was taken from parents and twins. Follow-up questionnaires have been administered at 6, 12, 18 and 36 months of age. All height and weight measures are self-reported by parents.

Up to 54 ART-conceived offspring and 101 NC offspring were included in this study (multiple births only). MUBICOS contributed results to the main analysis (ART vs. NC) to additional analysis stratified by sex, and IVF/ICSI (for IVF only), for height, weight, BMI. Data were available for most study confounders (maternal age, BMI, smoking, education, parity and offspring sex and age at outcome assessment), except ethnicity.

**22. Nascita e INFanzia: gli Effetti dell'Ambiente (NINFEA)**

NINFEA study is an Internet-based birth cohort established in 2005 in Italy (http://www.progettoninfea.it)^27^. A baseline questionnaire on general health and exposures before and during pregnancy is completed by mothers at enrolment, which may occur at any time during pregnancy. During the period 2005-2016 around 7,500 mothers were recruited. Further follow-up information was obtained with repeated questionnaires completed 6 and 18 months after delivery and when children turn 4, 7, 10 and 13 years. At each follow-up mothers reported their child current weight and height measurements, and if able to recall or retrieve from baby books or child health records retrospective measurements at pre-defined ages (3 months at 6-month, 12 months at 18-month, and 5 and 6 years at 7-year follow-up). Additional information on whether the measurements were recalled or taken from baby books is available.

Information on exposures was retrieved from the baseline questionnaire completed during pregnancy where mothers reported whether the pregnancy was planned or not, and in the case of affirmative response the following information was collected: i) number of months since she begun trying and became pregnant, ii) if she used any ART treatment, and iii) the type of ART as a checkbox (ovulation induction, intrauterine insemination, gamete intrafallopian transfer, in vitro fertilization, intra-cytoplasmic sperm injection, other technique). Information on the following confounding variables was collected in the baseline and 6-month follow-up questionnaires: maternal age (continuous), maternal BMI (continuous, derived from maternal pre-pregnancy weight and height reported at enrolment); smoking during pregnancy (yes vs. no, defined as any maternal smoking during pregnancy, independently whether sustained or not), educational level (low — primary school or less, medium—secondary school, and high—university degree), maternal country of birth as a proxy for ethnicity (born in Italy vs. born outside Italy, with more than 95% of mothers born in Italy), and parity (nulliparous vs. multiparous, based on the number of previous livebirths and stillbirths).

Up to 253 ART-conceived offspring and 4,990 NC offspring were included in this study. NINFEA contributed results to the main analysis (ART vs. NC), and to additional analyses stratified by sex, sub-fertility, and IVF/ICSI, for height, weight, and BMI. Data were available for all study confounders (maternal age, BMI, smoking, education, ethnicity, parity and offspring sex and age at outcome assessment).

**23. Norwegian Mother, Father and Child Cohort Study (MoBa)**

MoBa is a nationwide, pregnancy cohort comprising family triads (mother-father-offspring) who are followed longitudinally. All pregnant women in Norway who were able to read Norwegian were eligible to participate. The first child was born in 1999 and the last in 2009^28,29^. Extensive longitudinal data were collected using nine questionnaires: three during pregnancy, and then follow-up questionnaires when the children were 6 months, 18 months, 36 months, 5 years, 7 years and 8 years of age. Data collected include general background and health information, including diet and lifestyle, a semi-quantitative food frequency questionnaire, information on birth and pregnancy outcomes, and on several aspects of child nutrition and development, as well as the physical and mental health of both mother and child. MoBa is linked to the Medical Birth Registry of Norway, which provides standardized information about the health of the mother during pregnancy, other essential medical information related to the pregnancy and birth, and standard post-natal measures of the child. The Medical Birth Registry (MBRN) is a national health registry containing information about all births in Norway.

Up to 2,097 ART-conceived offspring and 77,210 NC offspring were included in this study (multiple birth included). MoBa contributed results to the main analysis (ART vs. NC) to all additional analysis, for height, weight, BMI. Data were available for most study confounders (maternal age, BMI, smoking, education, parity, and offspring sex and age at outcome assessment), but not ethnicity.

**24. Piccolipiù**

Piccolipiù is a prospective birth cohort study of 3,358 children born in selected maternal units located in five Italian cities (Florence, Rome, Trieste, Turin, and Viareggio) between 2011-2015. Piccolipiù study recruited singleton pregnant women aged at least 18 years old and giving birth in one of the selected maternity units. Mothers were recontacted when the child was 6, 12, 24, 48 months and 6 years old for follow-up questionnaires^30^.

Up to 86 ART-conceived offspring and 2,479 NC offspring were included in this study. Piccolipiù contributed results to the main analysis (ART vs. NC) to additional analysis stratified by sex, and sub-fertility, for height, weight, BMI, and waist circumference. Data were available for all study confounders (maternal age, BMI, smoking, education, ethnicity, parity and offspring sex and age at outcome assessment).

**25. Southampton Women's Survey (SWS)**

SWS is a population-based prospective birth cohort study of 12 583, initially non-pregnant, women aged 20–34 years, living in the city of Southampton, UK^31^. Assessments of lifestyle, diet and anthropometry were done at study entry in 1998–2002. Women who subsequently became pregnant with singleton pregnancies were followed up during pregnancy; and their offspring have been studied in infancy and childhood. Research nurses collected all anthropometric measurements on offspring and DXA scans were performed at various ages to determine body fat % and fat mass index Information on ART was obtained at the time of the first scan by questioning the mother.

Up to 36 ART-conceived offspring and 2,554 NC offspring were included in this study (singleton births only). SWS contributed results to the main analysis (ART vs. NC) to additional analysis stratified by sex, for all study outcomes (i.e., height, weight, BMI, waist circumference, body fat % and fat mass index). Data were available for all study confounders (maternal age, BMI, smoking, education, ethnicity, parity and offspring sex and age at outcome assessment).

**26. The Trøndelag Health Study (HUNT)**

The Trøndelag Health Study (HUNT) is a population-based study where all adult residents of the Nord-Trøndelag region, Norway have been invited to repeated surveys since the 1980s. Since the 1990s, all adolescents (aged 13-19 years) in the region have also been invited (the Young-HUNT Study)^32,33^. The participants have consented to data linkage to health registries, such as the Medical Birth Registry of Norway (MBRN), which includes information on virtually all births in Norway since 1967. In this study, we included participants from the Young-HUNT1 (1995-97), Young-HUNT2 (1999-2000) and Young-HUNT3 (2006-08) surveys, which included clinical measurements of height, weight and waist and hip circumferences. Information on mode of conception was obtained through linkage to information from the MBRN.

Up to 121 ART-conceived offspring and 9,711 NC offspring were included in this study (including multiple births). HUNT contributed results to the main analysis (ART vs. NC) to additional analysis stratified by sex, IVF/ICSI, and ET/FET, for height, weight, BMI, and waist circumference. Data were available for some study confounders (maternal age, parity and offspring sex and age at outcome assessment), but not for maternal BMI, smoking, education, or ethnicity.

### **eReferences**

1.	Tough SC, McDonald SW, Collisson BA, et al. Cohort Profile: The All Our Babies pregnancy cohort (AOB). *Int J Epidemiol* 2017; **46**(5): 1389-90k.

2.	van Eijsden M, Vrijkotte TG, Gemke RJ, van der Wal MF. Cohort profile: the Amsterdam Born Children and their Development (ABCD) study. *Int J Epidemiol* 2011; **40**(5): 1176-86.

3.	Boyd A, Golding J, Macleod J, et al. Cohort Profile: the 'children of the 90s'--the index offspring of the Avon Longitudinal Study of Parents and Children. *Int J Epidemiol* 2013; **42**(1): 111-27.

4.	Fraser A, Macdonald-Wallis C, Tilling K, et al. Cohort Profile: the Avon Longitudinal Study of Parents and Children: ALSPAC mothers cohort. *Int J Epidemiol* 2013; **42**(1): 97-110.

5.	Northstone K, Lewcock M, Groom A, et al. The Avon Longitudinal Study of Parents and Children (ALSPAC): an update on the enrolled sample of index children in 2019. *Wellcome open research* 2019; **4**: 51-.

6.	O'Donovan SM, Murray DM, Hourihane JO, Kenny LC, Irvine AD, Kiely M. Cohort profile: The Cork BASELINE Birth Cohort Study: Babies after SCOPE: Evaluating the Longitudinal Impact on Neurological and Nutritional Endpoints. *Int J Epidemiol* 2015; **44**(3): 764-75.

7.	Vuillermin P, Saffery R, Allen KJ, et al. Cohort Profile: The Barwon Infant Study. *Int J Epidemiol* 2015; **44**(4): 1148-60.

8.	Qiu X, Lu J-H, He J-R, et al. The Born in Guangzhou Cohort Study (BIGCS). *European Journal of Epidemiology* 2017; **32**(4): 337-46.

9.	Lewis S, Kennedy J, Burgner D, et al. Clinical review of 24-35 year olds conceived with and without in vitro fertilization: study protocol. *Reprod Health* 2017; **14**(1): 117-.

10.	Olsen J, Melbye M, Olsen SF, et al. The Danish National Birth Cohort - its background, structure and aim. *Scandinavian Journal of Public Health* 2001; **29**(4): 300-7.

11.	Heude B, Forhan A, Slama R, et al. Cohort Profile: The EDEN mother-child cohort on the prenatal and early postnatal determinants of child health and development. *International Journal of Epidemiology* 2015; **45**(2): 353-63.

12.	Charles MA, Thierry X, Lanoe J-L, et al. Cohort Profile: The French national cohort of children (ELFE): birth to 5 years. *International Journal of Epidemiology* 2019; **49**(2): 368-9j.

13.	Grote V, Theurich M, Luque V, et al. Complementary Feeding, Infant Growth, and Obesity Risk: Timing, Composition, and Mode of Feeding. *Nestle Nutr Inst Workshop Ser* 2018; **89**: 93-103.

14.	Weber M, Grote V, Closa-Monasterolo R, et al. Lower protein content in infant formula reduces BMI and obesity risk at school age: follow-up of a randomized trial. *The American journal of clinical nutrition* 2014; **99**(5): 1041-51.

15.	Koletzko B, von Kries R, Closa R, et al. Lower protein in infant formula is associated with lower weight up to age 2 y: a randomized clinical trial. *The American journal of clinical nutrition* 2009; **89**(6): 1836-45.

16.	Totzauer M, Luque V, Escribano J, et al. Effect of Lower Versus Higher Protein Content in Infant Formula Through the First Year on Body Composition from 1 to 6 Years: Follow-Up of a Randomized Clinical Trial. *Obesity (Silver Spring)* 2018; **26**(7): 1203-10.

17.	Porta D, Fantini MP. Prospective cohort studies of newborns in Italy to evaluate the role of environmental and genetic characteristics on common childhood disorders. *Italian Journal of Pediatrics* 2006; **32**: 350-7.

18.	Jaddoe VW, Mackenbach JP, Moll HA, et al. The Generation R Study: Design and cohort profile. *Eur J Epidemiol* 2006; **21**(6): 475-84.

19.	Kooijman MN, Kruithof CJ, van Duijn CM, et al. The Generation R Study: design and cohort update 2017. *European journal of epidemiology* 2016; **31**(12): 1243-64.

20.	Larsen PS, Kamper-Jørgensen M, Adamson A, et al. Pregnancy and birth cohort resources in europe: a large opportunity for aetiological child health research. *Paediatric and perinatal epidemiology* 2013; **27**(4): 393-414.

21.	Gallagher AL, Galvin R, Robinson K, Murphy C-A, Conway PF, Perry A. The characteristics, life circumstances and self-concept of 13 year olds with and without disabilities in Ireland: A secondary analysis of the Growing Up in Ireland (GUI) study. *PloS one* 2020; **15**(3): e0229599-e.

22.	Morton SMB, Atatoa Carr PE, Grant CC, et al. Cohort Profile: Growing Up in New Zealand. *International Journal of Epidemiology* 2012; **42**(1): 65-75.

23.	Soh S-E, Tint MT, Gluckman PD, et al. Cohort Profile: Growing Up in Singapore Towards healthy Outcomes (GUSTO) birth cohort study. *International Journal of Epidemiology* 2013; **43**(5): 1401-9.

24.	Moschonis G, Kalliora AC, Costarelli V, et al. Identification of lifestyle patterns associated with obesity and fat mass in children: the Healthy Growth Study. *Public Health Nutr* 2014; **17**(3): 614-24.

25.	Brescianini S, Fagnani C, Toccaceli V, et al. An update on the Italian Twin Register: advances in cohort recruitment, project building and network development. *Twin Res Hum Genet* 2013; **16**(1): 190-6.

26.	Connelly R, Platt L. Cohort Profile: UK Millennium Cohort Study (MCS). *International Journal of Epidemiology* 2014; **43**(6): 1719-25.

27.	Richiardi L, Baussano I, Vizzini L, Douwes J, Pearce N, Merletti F. Feasibility of recruiting a birth cohort through the Internet: the experience of the NINFEA cohort. *Eur J Epidemiol* 2007; **22**(12): 831-7.

28.	Magnus P, Birke C, Vejrup K, et al. Cohort Profile Update: The Norwegian Mother and Child Cohort Study (MoBa). *Int J Epidemiol* 2016; **45**(2): 382-8.

29.	Magnus P, Irgens LM, Haug K, Nystad W, Skjaerven R, Stoltenberg C. Cohort profile: the Norwegian Mother and Child Cohort Study (MoBa). *Int J Epidemiol* 2006; **35**(5): 1146-50.

30.	Farchi S, Forastiere F, Vecchi Brumatti L, et al. Piccolipiù, a multicenter birth cohort in Italy: protocol of the study. *BMC Pediatrics* 2014; **14**(1): 36.

31.	Inskip HM, Godfrey KM, Robinson SM, Law CM, Barker DJ, Cooper C. Cohort profile: The Southampton Women's Survey. *Int J Epidemiol* 2006; **35**(1): 42-8.

32.	Krokstad S, Langhammer A, Hveem K, et al. Cohort Profile: The HUNT Study, Norway. *International Journal of Epidemiology* 2012; **42**(4): 968-77.

33.	Holmen TL, Bratberg G, Krokstad S, et al. Cohort profile of the Young-HUNT Study, Norway: A population-based study of adolescents. *International Journal of Epidemiology* 2013; **43**(2): 536-44.

### Ethics approvals

| Cohort name | Ethic approval description |
| --- | --- |
| AOF | This study was approved by the Child Health Research Office and the Conjoint Health Research Ethics Board of the Faculties of Medicine, Nursing, and Kinesiology, University of Calgary, and the Affiliated Teaching Institutions (Ethics ID 20821 and 22821). Participants provided consent at the time of recruitment and were provided copies of the consent form for their records |
| ABCD | Approval for the ABCD study was obtained from the Central Committee on Research involving Human Subjects in the Netherlands, the Medical Ethical Committees of the participating hospitals, and from the Registration Committee of the Municipality of Amsterdam. Written informed consent was obtained from all participating mothers. |
| ALSPAC | Ethical approval for the study was obtained from the ALSPAC Ethics and Law Committee and the Local Research Ethics Committees. Informed consent for the use of data collected via questionnaires and clinics was obtained from participants following the recommendations of the ALSPAC Ethics and Law Committee at the time. At age 18, study children were sent 'fair processing' materials describing ALSPAC’s intended use of their health and administrative records and were given clear means to consent or object via a written form. Data were not extracted for participants who objected, or who were not sent fair processing materials. Ethical approval for the study was obtained from the ALSPAC Law and Ethics committee and local research ethics committees (NHS Haydock REC: 10/H1010/70). |
| BASELINE | Research objectives and measurements in this birth cohort were conducted according to the guidelines laid down in the Declaration of Helsinki and all procedures were approved by the Clinical Research Ethics Committee of the Cork Teaching Hospitals, [ref ECM5(9) 01/07/ 2008]. Families provided written informed consent at 20 weeks’ gestation or at birth to participate in BASELINE follow-up. |
| BIS | Ethics approval was obtained from the Barwon Health Human Research Ethics Committee (10/24). All mothers provided written informed consent. |
| BIGCS | The study protocol was approved by the institutional ethics committee of the Guangzhou Women and Children’s Medical Center. Written informed consent was provided by all participants. |
| CHART | The study was approved by the Royal Children's Hospital Human Research Ethics Committee, and all study participants provided consent to take part in the study. |
| DNBC | The DNBC complies with the Declaration of Helsinki and was approved by the Danish National Committee on Biomedical Research Ethics. Informed consent was obtained from participants upon enrolment. |
| EDEN | The study received approval from the ethics committee (CCPPRB) of Kremlin Bicêtre on 12 December 2002 and from CNIL (Commission Nationale Informatique et Liberté), the French data privacy institution. All subjects gave their informed consent for inclusion before they participated in the study. Consent for the child was obtained from both parents after the child's birth. |
| ELFE | Ethical approvals for data collection in maternity units and for each data collection wave during follow-up were obtained from the national advisory committee on information processing in health research (CCTIRS: Comité Consultatif sur le Traitement de l’Information en matière de Recherche dans le domaine de la Santé), the national data protection authority (CNIL: Comission Nationale Informatique et Liberté) and, in case of invasive data collection such as biological sampling, the committee for protection of persons engaged in research (CPP: Comité de Protection des Personnes). The ELFE study was also approved by the national committee for statistical information (CNIS: Conseil National de l’Information Statistique). Informed consent was signed by the parents or the mother alone, with the father being informed of his right to deny consent for participation |
| CHOP | The study was approved by the ethics committees of all study centres. Written informed parental consent was obtained for each infant. All research was conducted in accordance with the Declaration of Helsinki. |
| GASPII | The protocol of the study has been approved by the Ethics committees of the Università Cattolica del Sacro Cuore, Rome, and all study participants provided consent to take part in the study. |
| Gen-R | The study has been approved by the Medical Ethical Committee of the Erasmus MC, University Medical Center in Rotterdam (MEC-2012-165-NL40020.078.12). Written informed consent was obtained from the parents or legal representatives of the children. Even with consent of the parents, when the child is not willing to participate actively, no measurements are performed. |
| G21 | The Ethics Committee of Hospital de São João, and of the Institute of Public Health of the University of Porto approved the study protocols. The study complies with the Ethical Principles expressed in the Helsinki Declaration and with the national legislation and was registered with the Portuguese Authority for Data Protection. In all evaluations, participants were informed about the purposes and design of the study, as well as the potential discomfort caused by participation. Signed informed consent was obtained from all parents or legal guardians, and oral assent was obtained from children at each evaluation. |
| GUI | The GUI study received independent ethics approval from a Research Ethics Committee convened by the Department of Health and Children. Written informed consent was obtained from parents or guardians. All methods were performed in accordance with the relevant guidelines and regulations. |
| GUiNZ | Ethical approval for GUiNZ was received from the Ministry of Health Northern Y Regional Ethics Committee (NTY/08/06/055). Written informed consent was obtained from all participating mothers at recruitment and confirmed at each subsequent interview. |
| GUSTO | Study protocols following the principles of the Declaration of Helsinki and were approved by the respective ethics committees for two hospitals: National Healthcare Group Domain Specific Review Board (NUH) and SingHealth Centralized Institutional Review Board (KKH). All participants in this study provided informed consent to participate and contribute their data to publications. The GUSTO study is registered under study ID: NCT01174875 (clinicaltrials.gov) which broadly covers investigations of parental and gestational influences on child health. |
| HGS | Approval to conduct the study was granted by the Greek Ministry of National Education and the Ethics Committee of Harokopio University of Athens, and the study was conducted in accordance with the ethical standards specified in the 1964 Declaration of Helsinki. Parents who agreed to the participation of their children in the study had to sign the consent form and provide their contact details. |
| ITR | The study was approved by the ethics committee of Istituto Superiore di Sanità (prot. Number CE-ISS 05-113). All included twins gave their consent to participate in the studies proposed by the ITR research group. |
| MCS | Ethical approval for the Millennium Cohort Study was granted from the multi-centre research ethics committee. Following ethical approval for the study from an NHS Research Ethnics Committee (MREC), informed consent was obtained from parents, as well as from the children themselves as they grew up. |
| MUBICOS | The study was approved by the ethics committee of Istituto Superiore di Sanità (prot. Number CE-ISS 09-281). All included twins gave their consent to participate in the studies proposed by the ITR research group. |
| NINFEA | The Ethical Committee of the San Giovanni Battista Hospital and CTO/CRF/Maria Adelaide Hospital of Turin approved the NINFEA study (approval N. 0048362, and subsequent amendments). Informed consent was obtained from all the participants at enrolment and at each follow-up. |
| MoBa | The establishment and data collection in MoBa was previously based on a license from the Norwegian Data protection agency and approval from The Regional Committee for Medical Research Ethics, and it is now based on regulations related to the Norwegian Health Registry Act. MoBa is conducted according to the guidelines laid down in the declaration of Helsinki, and written informed consent was obtained from all participants. A detailed protocol of the study including the consent can be found elsewhere ([http://www.fhi.no/morogbarn](about:blank)). |
| Piccolipiù | The protocol of the study has been approved by the Ethics committees of the Local Health Unit Roma E (management centre), of the Istituto Superiore di Sanità (National Institute of Public Health) and of each local centre. Standard procedures for the protection of confidential individual information were applied according to the Italian law. Consent forms for participation was signed by the mother and also by the father, when both legally responsible for the newborn. |
| SWS | SWS study was conducted according to the guidelines laid down in the Declaration of Helsinki and was approved by the Southampton and South West Hampshire Local Research Ethics Committee (06/Q1702/104). Written informed consent was obtained from all participating women and by a parent or guardian with parental responsibility on behalf of their children. |
| HUNT | The study is approved by the Regional Committee for Medical and Health Research Ethics and by the Norwegian Data Protection Authority, and all study participants gave consent to take part in the study. |

### **eTable 1. Overview of the participating cohorts**

| Cohort name | Cohort country | Birth years | Analysis sample size (% female) | Number of ART offspring included (%) | Offspring growth/adiposity outcome(s) included in analysis (number of repeat measurements) | Number of meta-analysis age groups contribution (and mean age/range of mean ages if included in >1 age group) |
| --- | --- | --- | --- | --- | --- | --- |
| AOF | AU | 2008-2011 | 1,804 (47.8) | 41 (2.3) | weight, height, BMI | 4 (1.1y to 5.2y) |
| ABCD | NL | 2003-2004 | 4,510 (55.5) | 61 (1.4) | weight, height, BMI, waist, bio, FMI | 10 (0.2y to 11.7y) |
| ALSPAC | UK | 1990-1992 | 8,652 (48.8) | 53 (0.6) | weight, height, BMI, waist, bio, FMI | 9 (0.1y to 24.5y) |
| BASELINE | IE | 2008-2011 | 1,051 (48.7) | 20 (1.9) | weight, height, BMI, waist, bio, FMI | 5 (0.2y to 5.1y) |
| BIS | AU | 2010-2013 | 708 (47.6) | 35 (4.9) | weight, height, BMI, bio | 2 (1.1y and 4.2y) |
| BIGCS | CH | 2012-present | 10,074 (47.7) | 349 (3.5) | weight, height, BMI | 4 (0.1y to 2.8y) |
| CHART | AU | 1982-1992 | 203 (60.6) | 130 (64.0) | weight, height, BMI, waist, bio, FMI | 1 (27.4y) |
| DNBC | DN | 1996-2003 | 36,380 (48.6) | 1,481 (4.1) | weight, height, BMI, waist | 4 (0.4y to 11.3y) |
| EDEN | FR | 2003-2006 | 1,348 (48.0) | 22 (1.6) | weight, height, BMI, waist, bio, FMI | 6 (0.3y to 5.7y) |
| ELFE | FR | 2011 | 9,941 (48.9) | 309 (3.1) | weight, height, BMI | 5 (0.3y to 3.0y) |
| CHOP | EU | 2002-2004 | 1,499 (50.0) | 20 (1.3) | weight, height, BMI, waist | 4 (0.1y to 1.0y) |
| GASPII | IT | 2003-2004 | 562 (48.8) | 8 (1.4) | weight, height, BMI, waist | 3 (1.4y to 7.8y) |
| Gen-R | NL | 2002-2006 | 4,307 (49.6) | 51 (1.2) | weight, height, BMI, waist, bio, FMI | 6 (0.1y to 9.8y) |
| G21 | PO | 2005-2006 | 4,756 (48.8) | 92 (1.9) | weight, height, BMI, waist, bio, FMI | 11 (0.2y to 10.2y) |
| GUI | IE | 2011 | 9,915 (48.8) | 173 (1.7) | weight, height, BMI | 3 (0.8y to 5.2y) |
| GUiNZ | NZ | 2009-2010 | 4,447 (48.9) | 173 (3.9) | weight, height, BMI, waist | 3 (2.0y to 8.6y) |
| GUSTO | SG | 2009-2010 | 905 (48.3) | 64 (7.1) | weight, height, BMI, waist, bio, FMI | 10 (0.1y to 6.1y) |
| HGS | GR | 2007-2009 | 2,245 (50.7) | 63 (2.8) | weight, height, BMI, waist, bio, FMI | 1 (11.2 years) |
| ITR | IT | 2003-2018 | 248 (49.2) | 54 (21.8) | weight, height, BMI | 7 (0.6y to 13.6y) |
| MCS | UK | 2000-2002 | 2,183 (48.0) | 30 (1.4) | weight, height, BMI, waist, bio, FMI | 5 (3.1y to 17.2y) |
| MUBICOS | IT | 2009-2015 | 155 (48.4) | 54 (34.8) | weight, height, BMI | 2 (1.1y and 3.0y) |
| NINFEA | IT | 2006-2017 | 5,260 (49.4) | 270 (5.1) | weight, height, BMI | 8 (0.3y to 10.2y) |
| MoBa | NO | 1998-2008 | 79,358 (49.1) | 2,148 (2.7) | weight, height, BMI | 10 (0.1y to 7.1y) |
| Piccolipiù | IT | 2011-2014 | 2,565 (48.3) | 86 (3.4) | weight, height, BMI, waist | 7 (0.1y to 4.4y) |
| SWS | UK | 1998-2005 | 2,589 (48.2) | 35 (1.4) | weight, height, BMI, waist, bio, FMI | 7 (0.5y to 9.2y) |
| HUNT | NO | 1984-2006 | 9,832 (49.8) | 121 (1.2) | weight, height, BMI, waist | 2 (15.0y to 18.1y) |
| Potentially eligible cohorts not included in analyses | | | | | | |
| BiB* | UK | 2007-2011 | 13,740 (49.3) | 7 | weight, height, BMI | - |
| Raine* | AU | 1989-1991 | around 5,000 | 5 | weight, height, BMI, waist, bio, FMI | - |
| PREDO* | FI | 2006-2010 | around 2,500 | - | weight, height, BMI | - |
| UBCoS* | SE | - | - | - | - | - |

For studies with repeat measures (and multiple outcomes), sample size and number of ART is shown for timepoint with the largest number of ART offspring. Thirty likely eligible cohorts were invited and all, except one, agreed to participate in this study. We had pre-specified that to be included cohorts should have data on at least 10 ART-conceived infants. Of those that agreed, all, except three, completed their analysis and were included. Two of the three cohorts informed us that they had fewer than 10 ART-conceived infants and the other one did not to respond to repeated requests to complete the analysis. The four excluded cohorts are indicated by *. The BiB (Born in Bradford: <https://borninbradford.nhs.uk/>) and Raine (The Western Australian Pregnancy Cohort: <https://rainestudy.org.au/>) studies did not participate because once they checked they reported that they had too few offspring conceived using ART according to our criterial of cohorts having to have at least 10 ART conceived infants. PREDO (Prediction and Prevention of Preeclampsia and Intrauterine Growth Restriction: <https://academic.oup.com/ije/article/46/5/1380/2622848>) initially agreed to contribute but unfortunately did not respond to subsequent requests to run the analysis and was excluded from the meta-analysis. UBCoS (Uppsala Birth Cohort Multigeneration Study: <https://www.chess.su.se/ubcosmg/>) did not respond to the initial invitations to participate in this study and was excluded.

### **eTable 2. Descriptive data on participants numbers and outcomes in each included cohort**

See excel document: eTable_2.xlsx

### **eFigure 1. Directed Acyclic Graph used to identify potential confounders**

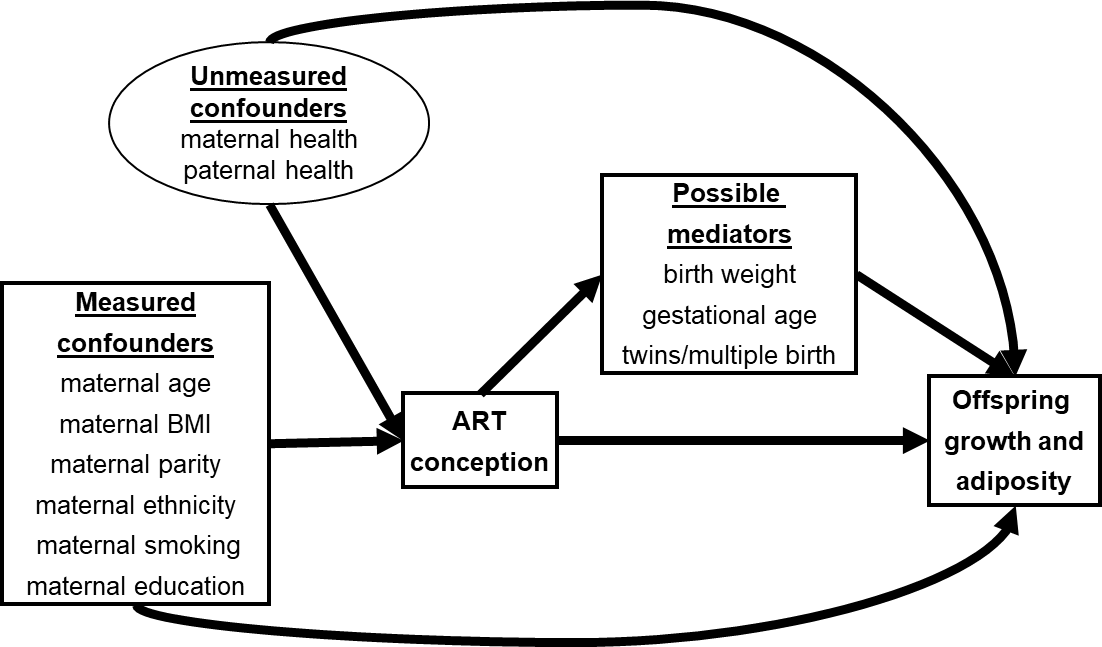

### **eFigure 2. Cohort-specific mean differences in length/height between ART-conceived and NC offspring**

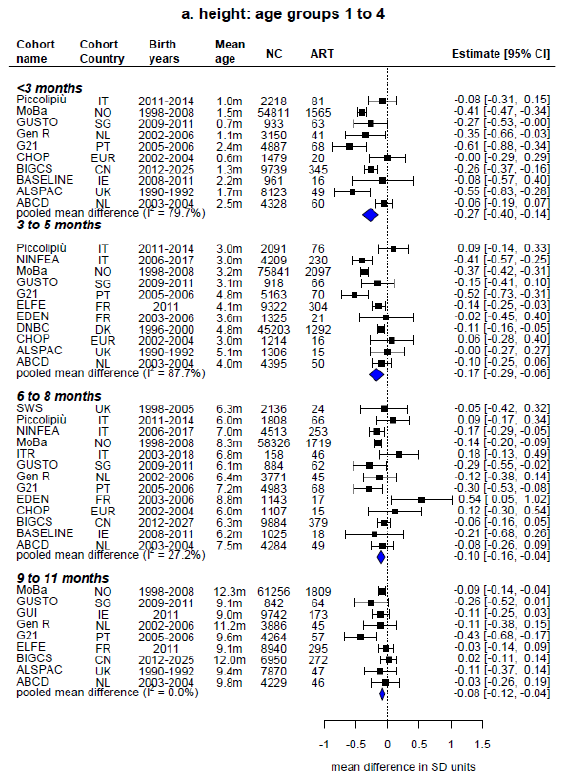

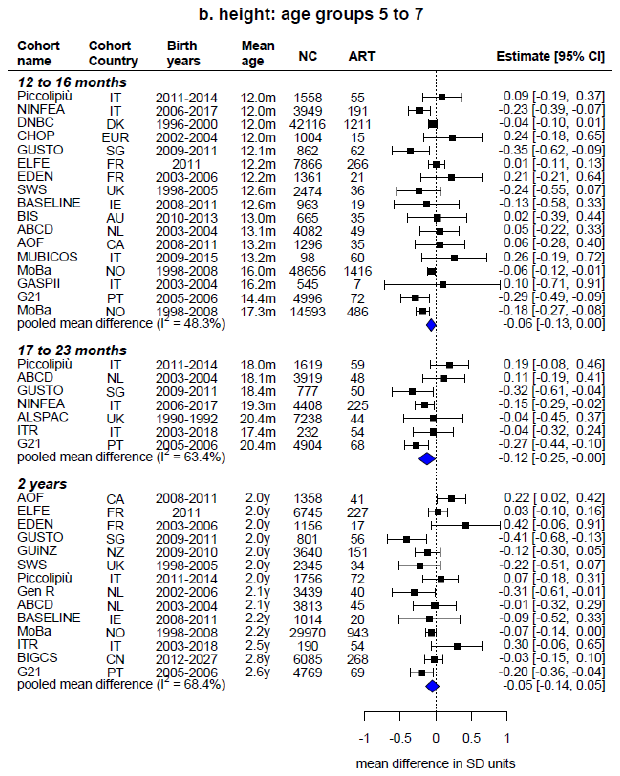

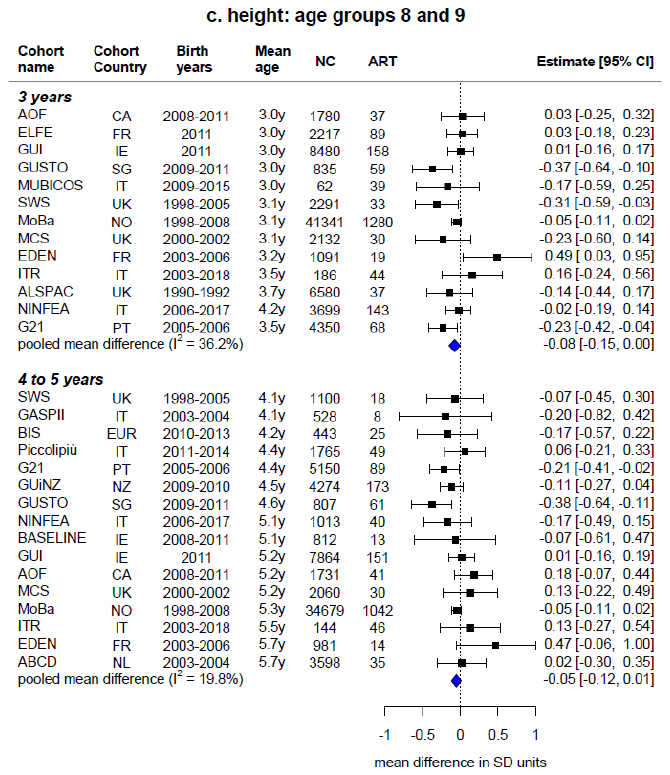

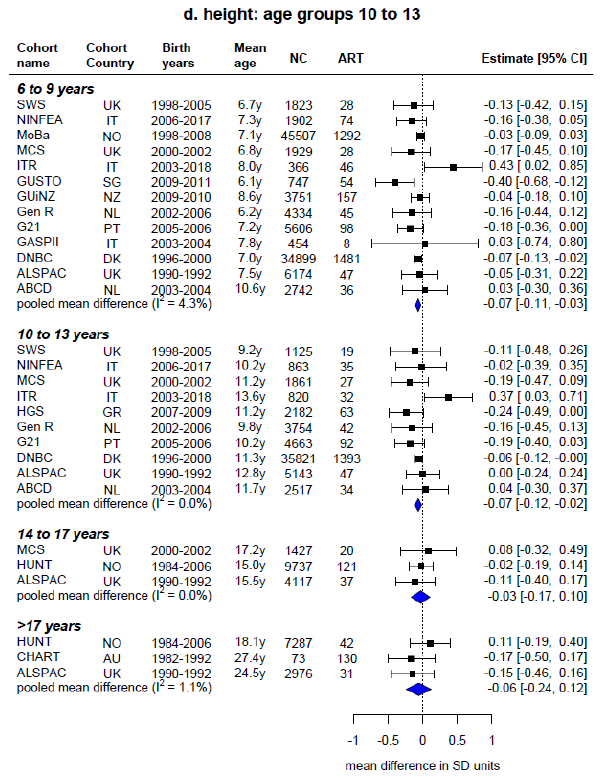

Estimates represent the cohort-specifc confounder-adjusted mean differences in SD units [and 95% confidence intervals] in length/height at each age group between ART-conceived and NC offspring (ART minus NC). Estimates were adjusted (as fully as possible) for maternal age, parity, BMI, smoking, education, ethnicity (or country of birth), plus offspring sex and age at outcome assessment. Cohorts are arranged by the offspring’s mean age at outcome assessment. Blue diamonds represent the pooled mean differences from random-effects meta-analyses. NC is the number of NC ofspring; ART is the number of ART-conceived offspring; I² represents the percentage of total variability due to between cohort heterogeneity.

### **eFigure 3. Cohort-specific mean differences in weight between ART-conceived and NC offspring**

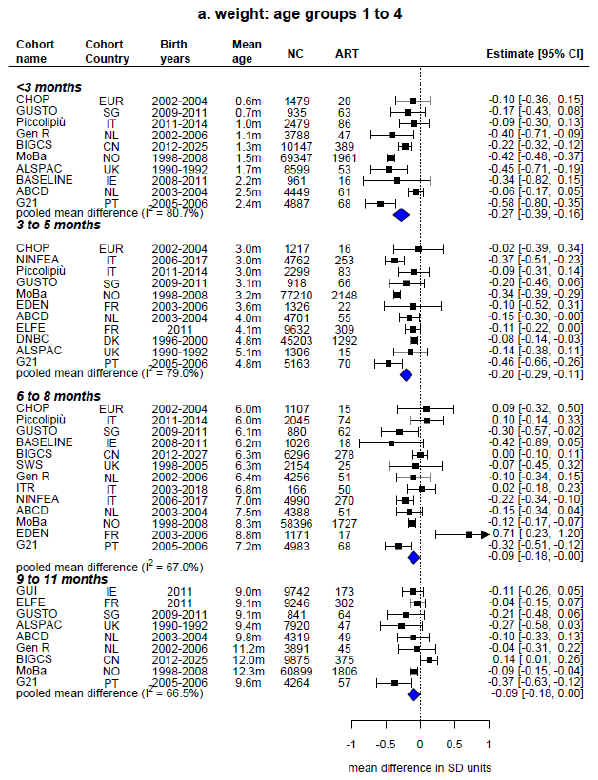

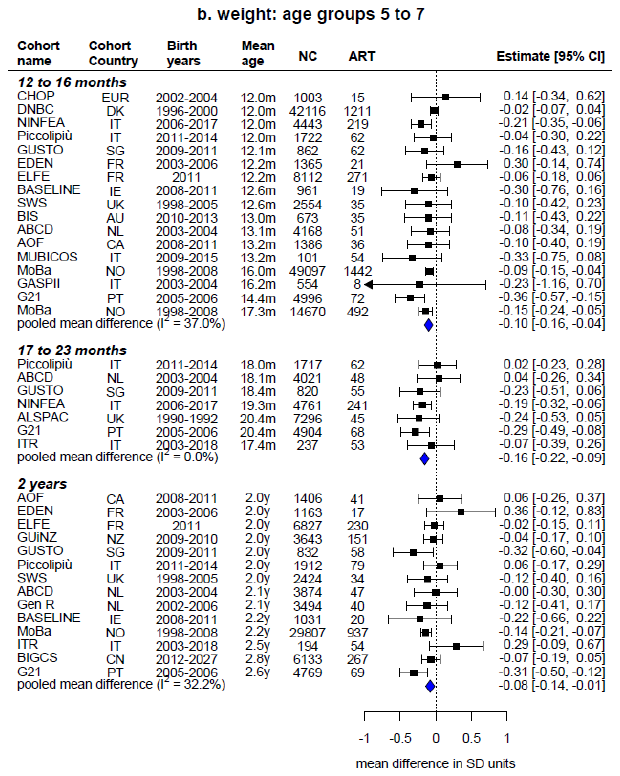

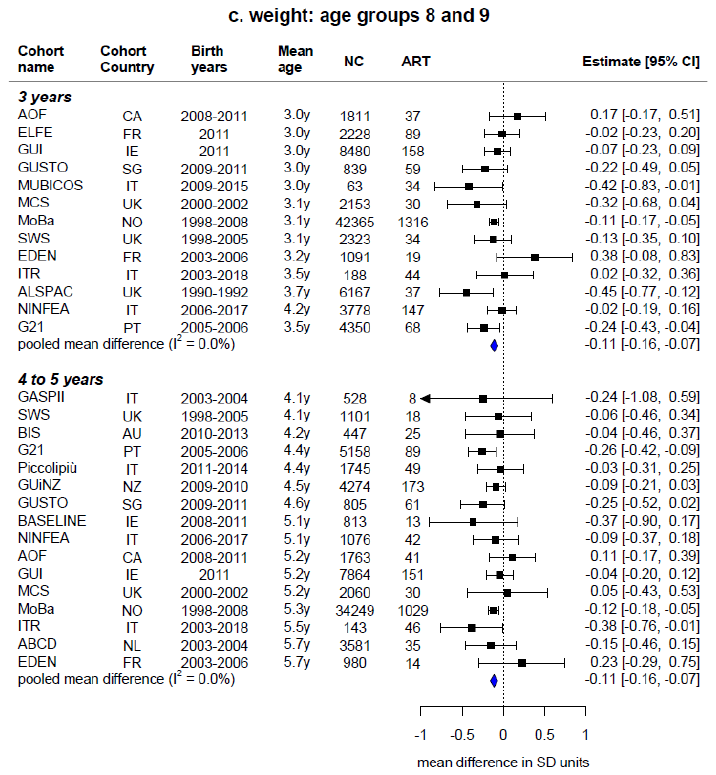

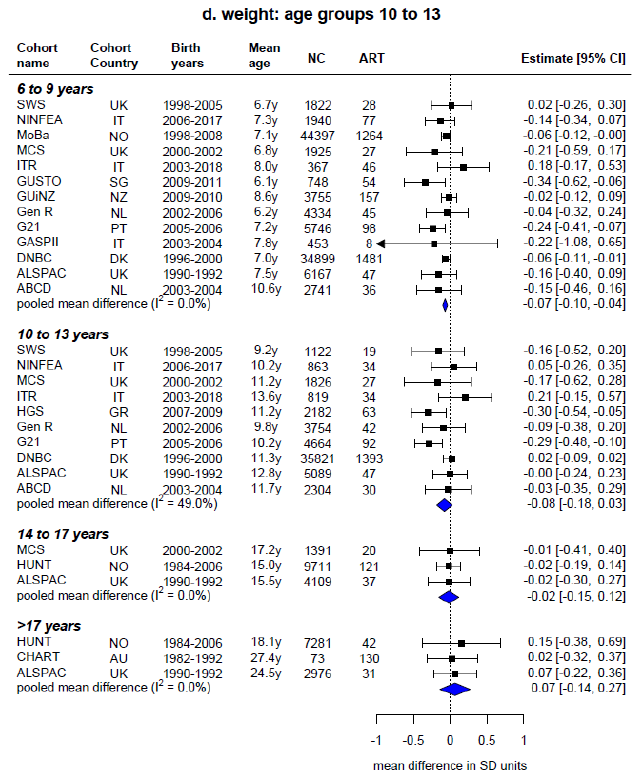

Estimates represent the cohort-specifc confounder-adjusted mean differences in SD units [and 95% confidence intervals] in weight at each age group between ART-conceived and NC offspring (ART minus NC). Estimates were adjusted (as fully as possible) for maternal age, parity, BMI, smoking, education, ethnicity (or country of birth), plus offspring sex and age at outcome assessment. Cohorts are arranged by the offspring’s mean age at outcome assessment. Blue diamonds represent pooled mean differences from random-effects meta-analyses. NC is the number of NC ofspring; ART is the number of ART-conceived offspring; I² represents the percentage of total variability due to between cohort heterogeneity.

### **eFigure 4. Cohort-specific mean differences in body mass index between ART-conceived and NC offspring**

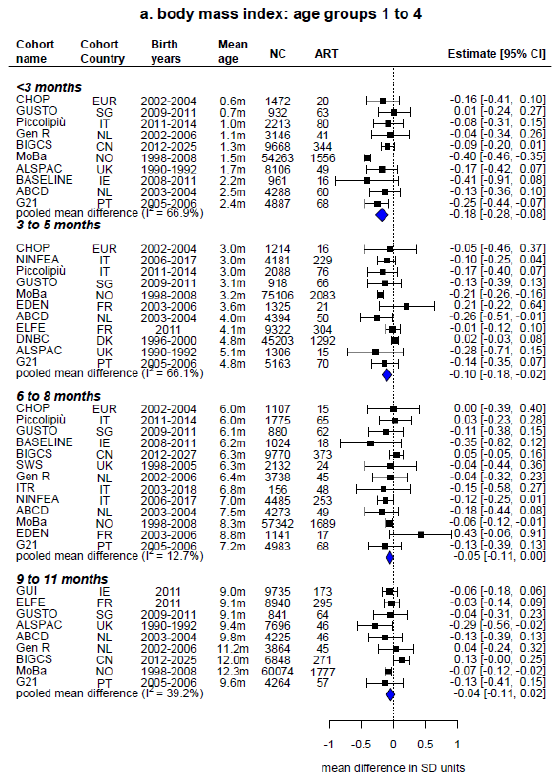

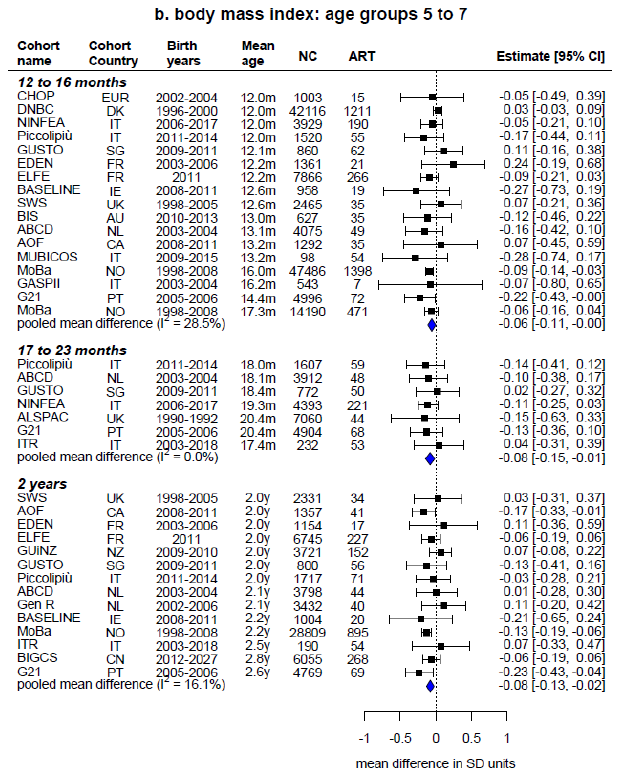

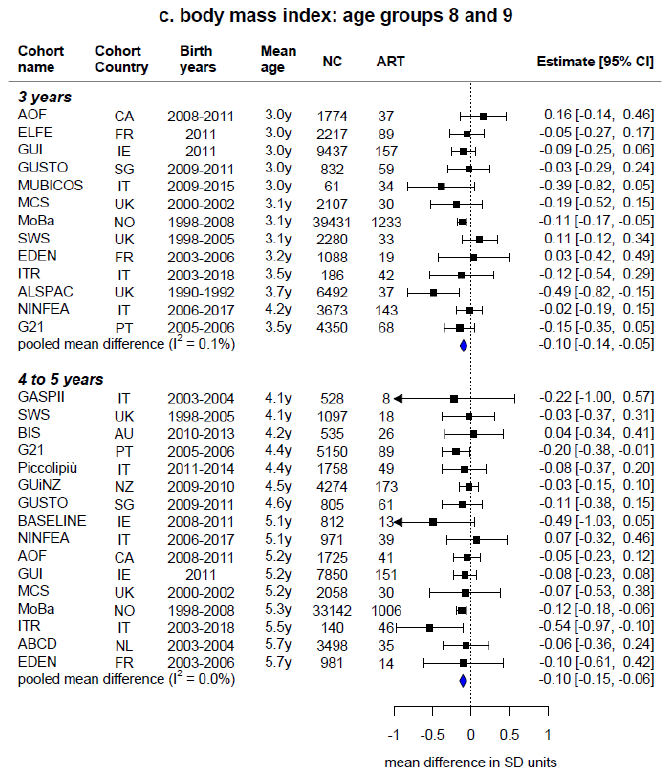

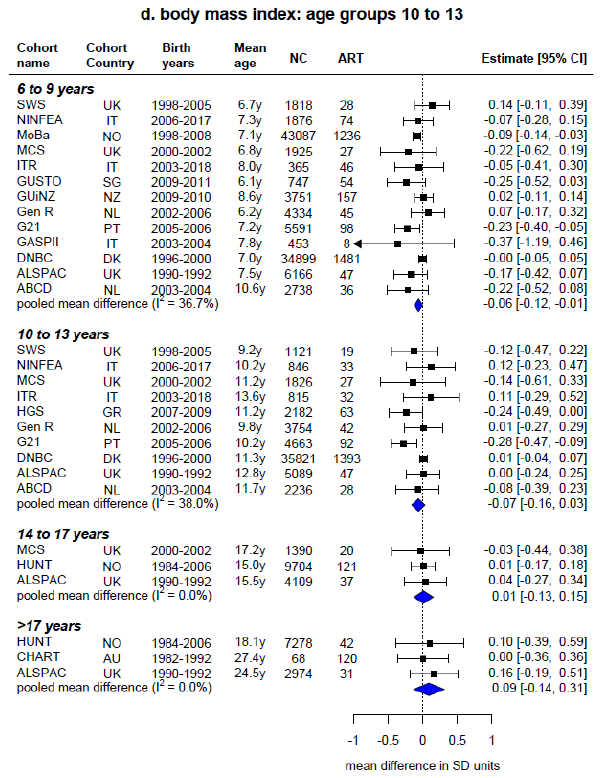

Estimates represent the cohort-specifc confounder-adjusted mean differences in SD units [and 95% confidence intervals] in body mass index at each age group between ART-conceived and NC offspring (ART minus NC). Estimates were adjusted (as fully as possible) for maternal age, parity, BMI, smoking, education, ethnicity (or country of birth), plus offspring sex and age at outcome assessment. Cohorts are arranged by the offspring’s mean age at outcome assessment. Blue diamonds represent the pooled mean differences from random-effects meta-analyses. NC is the number of NC ofspring; ART is the number of ART-conceived offspring; I² represents the percentage of total variability due to between cohort heterogeneity.

### **eFigure 5. Cohort-specific mean differences in waist circumference between ART-conceived and NC offspring**

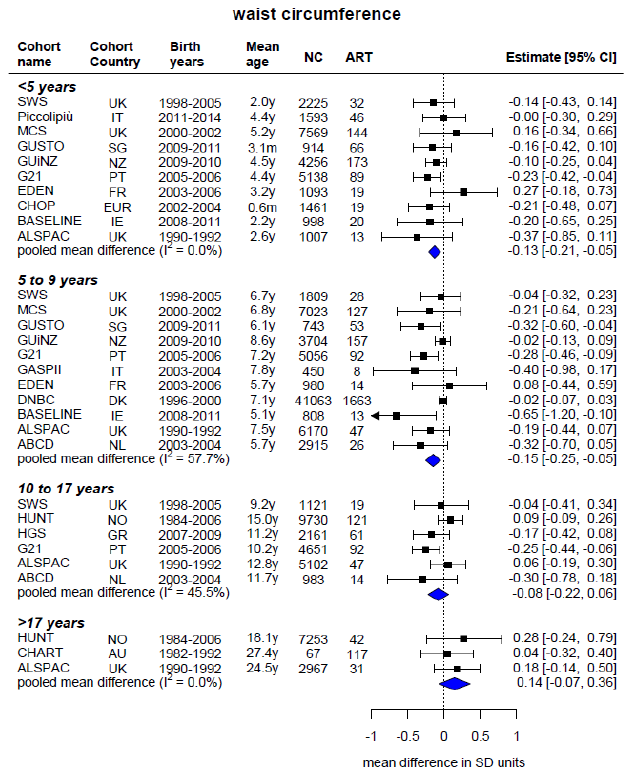

Estimates represent the cohort-specifc confounder-adjusted mean differences in SD units [and 95% confidence intervals] in waist circumference at each age group between ART-conceived and NC offspring (ART minus NC). Estimates were adjusted (as fully as possible) for maternal age, parity, BMI, smoking, education, ethnicity (or country of birth), plus offspring sex and age at outcome assessment. Cohorts are arranged by the offspring’s mean age at outcome assessment. Blue diamonds represent the pooled mean differences from random-effects meta-analyses. NC is the number of NC ofspring; ART is the number of ART-conceived offspring; I² represents the percentage of total variability due to between cohort heterogeneity.

### **eFigure 6. Cohort-specific mean differences in body fat % between ART-conceived and NC offspring**

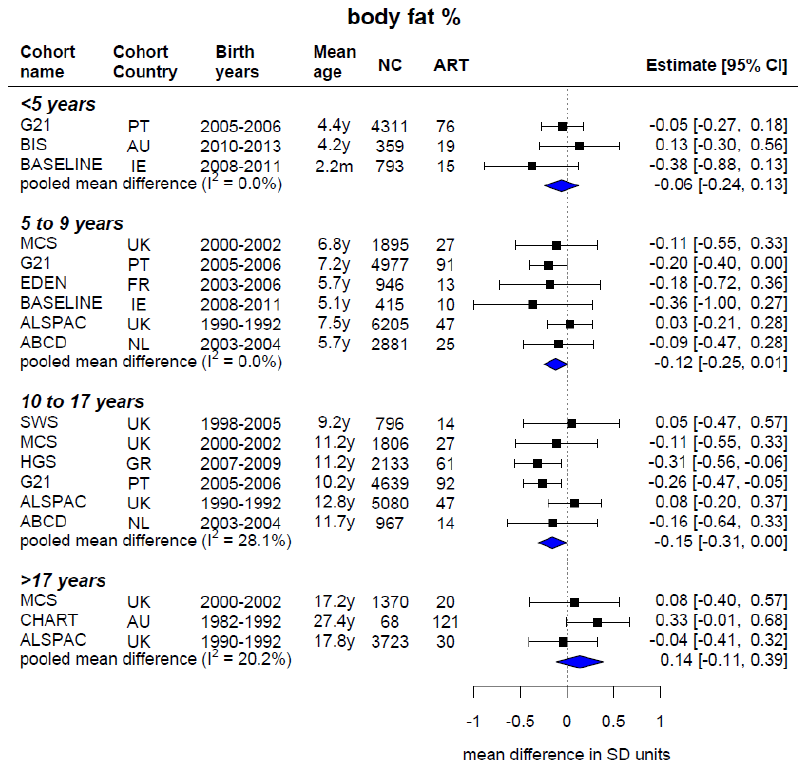

Estimates represent the cohort-specifc confounder-adjusted mean differences in SD units [and 95% confidence intervals] in body fat % at each age group between ART-conceived and NC offspring (ART minus NC). Estimates were adjusted (as fully as possible) for maternal age, parity, BMI, smoking, education, ethnicity (or country of birth), plus offspring sex and age at outcome assessment. Cohorts are arranged by the offspring’s mean age at outcome assessment. Blue diamonds represent the pooled mean differences from random-effects meta-analyses. NC is the number of NC ofspring; ART is the number of ART-conceived offspring; I² represents the percentage of total variability due to between cohort heterogeneity.

### **eFigure 7. Cohort-specific mean differences in fat mass index between ART-conceived and NC offspring**

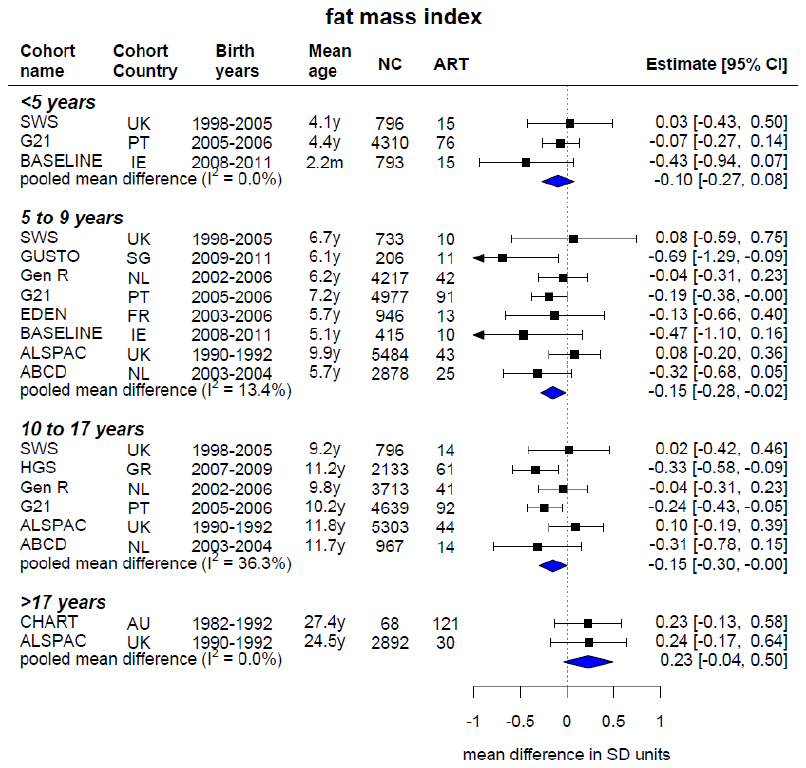

Estimates represent the cohort-specifc confounder-adjusted mean differences in SD units [and 95% confidence intervals] in fat mass index at each age group between ART-conceived and NC offspring (ART minus NC). Estimates were adjusted (as fully as possible) for maternal age, parity, BMI, smoking, education, ethnicity (or country of birth), plus offspring sex and age at outcome assessment. Cohorts are arranged by the offspring’s mean age at outcome assessment. Blue diamonds represent the pooled mean differences from random-effects meta-analyses. NC is the number of NC ofspring; ART is the number of ART-conceived offspring; I² represents the percentage of total variability due to between cohort heterogeneity.

### **eFigure 8. Mean difference in length/height, weight, and body mass index between ART-conceived and NC offspring at ages <3 months and 3-5 months, after leaving each cohort study out of the meta-analysis (to identify influential cohorts)**

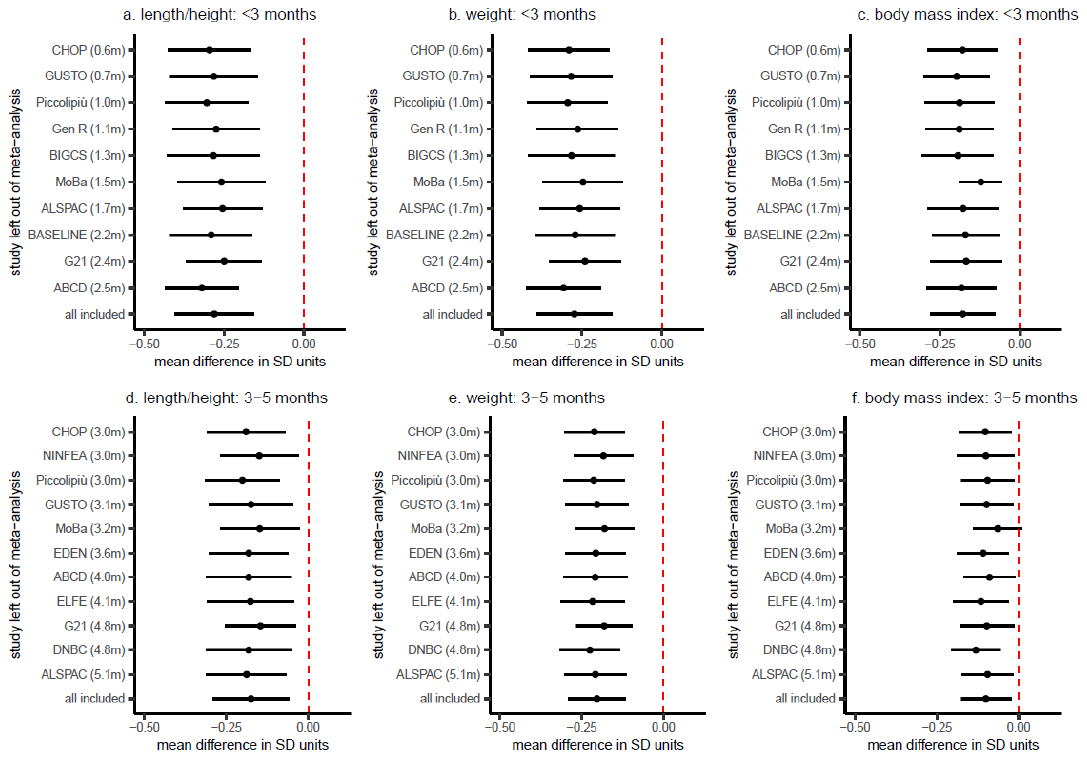

Estimates represent the confounder-adjusted pooled mean differences in SD units [and 95% confidence intervals] in length/height, weight, and body mass index at ages <3 months and 3-5 months between ART-conceived and NC offspring (ART minus NC), across all studies (bottom rows) and after refitting the meta-analysis models with each cohort study omitted in turn. Estimates were adjusted (as fully as possible) for maternal age, parity, BMI, smoking, education, ethnicity (or country of birth), plus offspring sex and age at outcome assessment. Cohorts arranged by offspring’s mean age at outcome assessment.

### **eFigure 9. Mean difference in growth and adiposity outcomes between ART-conceived and NC offspring, stratified by sex**

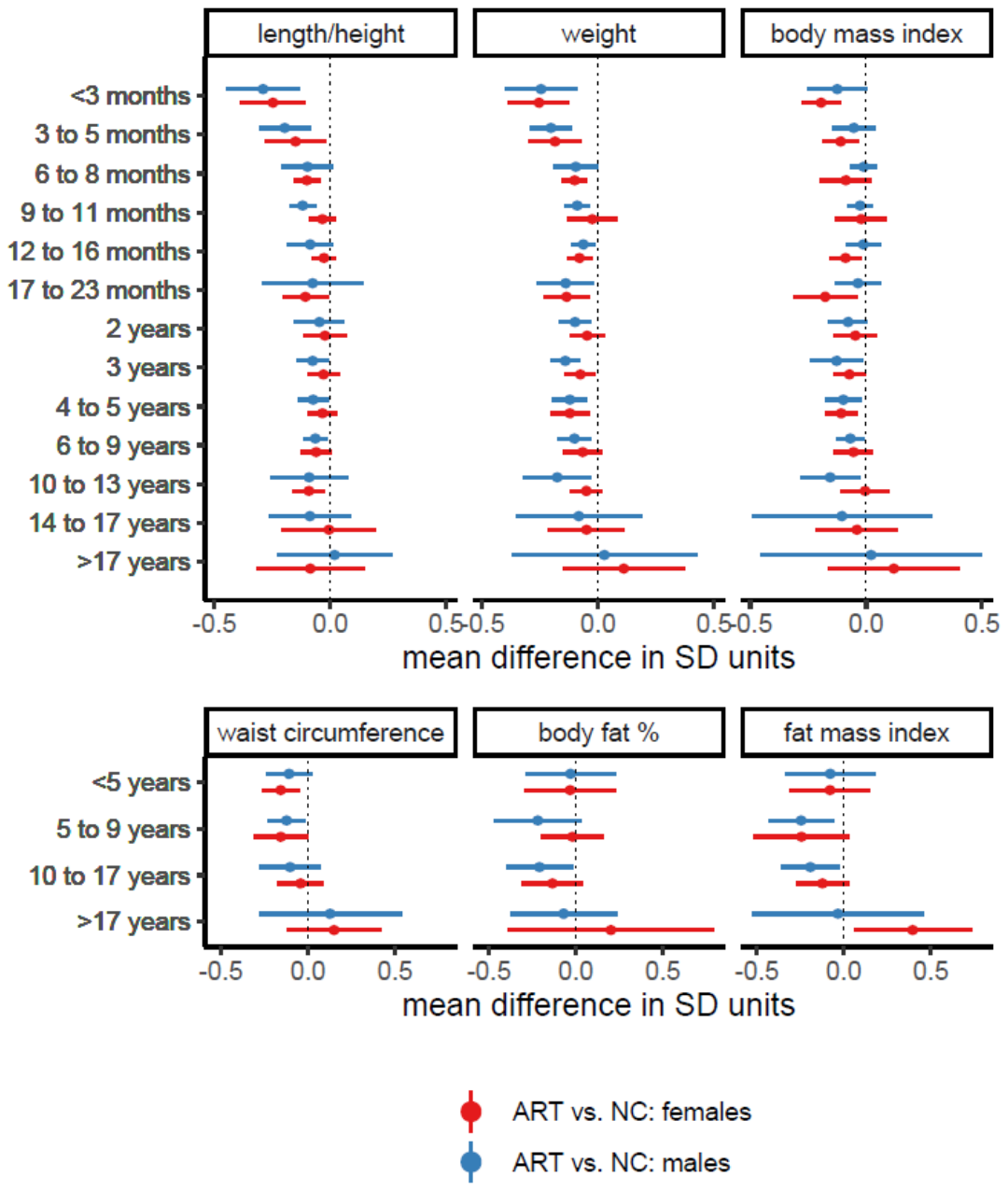

Estimates represent the confounder-adjusted pooled mean differences in SD units [and 95% confidence intervals] in growth and adiposity outcomes at each age group between ART-conceived and NC offspring (ART minus NC), seperately in females and males. Estimates were adjusted (as fully as possible) for maternal age, parity, BMI, smoking, education, ethnicity (or country of birth), plus offspring age at outcome assessment.

### **eFigure 10. Mean difference in growth and adiposity outcomes between ART-conceived and NC offspring, comparing results in all participants to singleton births only**

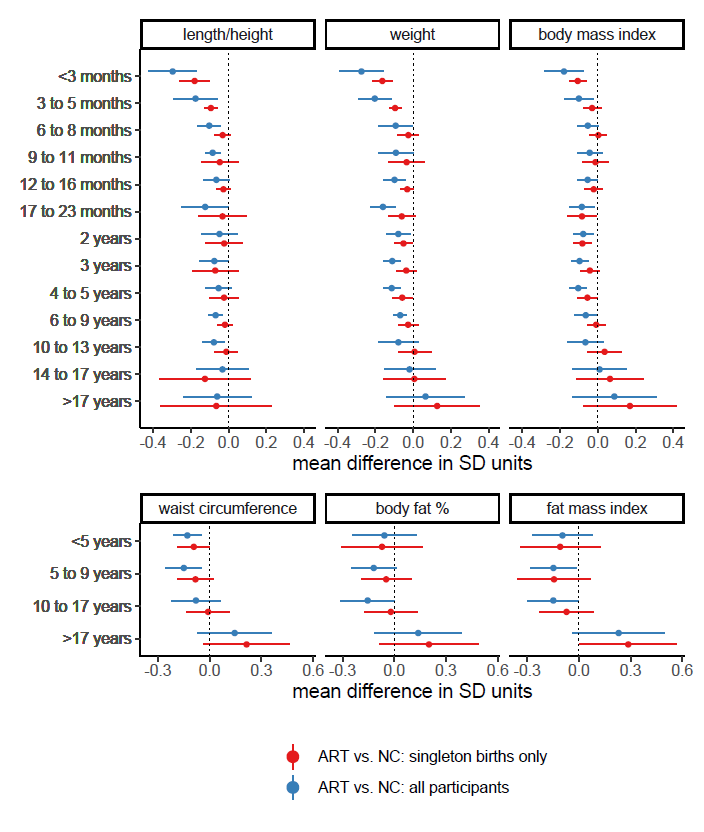

Estimates represent the confounder-adjusted pooled mean differences in SD units and 95% confidence intervals in growth and adiposity outcomes at each age group between ART-conceived and NC offspring (ART minus NC), comparing results in all participants (i.e., those presented in Figures 1-2) to singleton birth offspring. Cohort-specific estimates were adjusted (as fully as possible) for maternal age, parity, BMI, smoking, education, ethnicity (or country of birth), plus offspring sex and age at outcome assessment. Of the total 26 cohorts in this study, 15 cohorts included both singletons and multiple births, 9 cohorts included singletons only, and 2 cohorts included multiple births only.

### **eFigure 11. Mean difference in growth and adiposity outcomes between ART-conceived and NC offspring, after further adjustment for birth weight and gestational age**

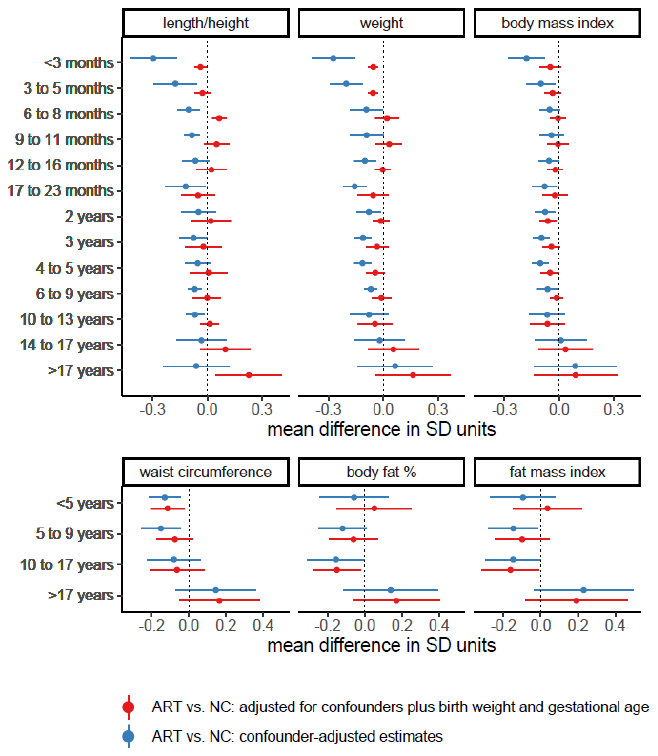

Estimates represent the confounder-adjusted pooled mean differences in SD units and 95% confidence intervals in growth and adiposity outcomes at each age group between ART-conceived and NC offspring (ART minus NC), before and after further adjustment for the potential mediators birthweight and gestational age. The confounder-adjusted estimates were adjusted (as fully as possible) for maternal age, parity, BMI, smoking, education, ethnicity (or country of birth), plus offspring sex and age at outcome assessment.

### **eFigure 12. Mean difference in length/height, weight, and body mass index between ART-conceived and NC offspring, separately for fresh and frozen-thawed embryo transfer, after further adjustment for birth weight and gestational age**

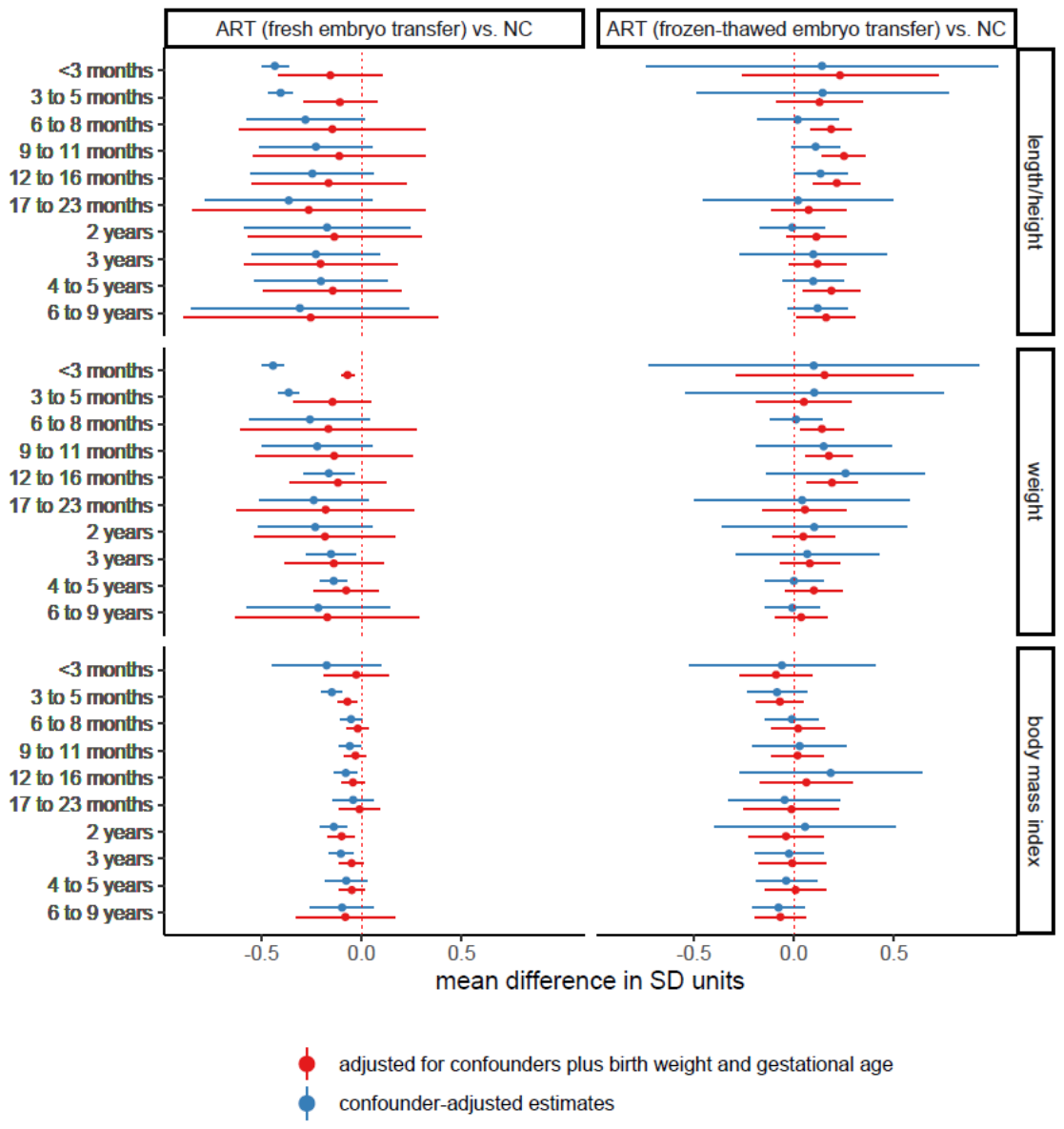

Estimates represent the confounder-adjusted pooled mean differences in SD units and 95% confidence intervals in length/height, weight, and body mass index at each age group between ART-conceived and NC offspring (ART minus NC), separately for fresh embryo transfer and frozen-thawed embryo transfer, before and after further adjustment for the potential mediators birthweight and gestational age. The confounder-adjusted estimates were adjusted (as fully as possible) for maternal age, parity, BMI, smoking, education, ethnicity (or country of birth), plus offspring sex and age at outcome assessment.

### **Cohort-specific acknowledgements and funding**

**All Our Families Study (AOF)**

The authors acknowledge the tremendous contribution and support of AOB/F participants and AOB/F team members. All Our Families is funded through Alberta Innovates Interdisciplinary Team Grant #200700595, the Alberta Children’s Hospital Foundation, the Canadian Institutes of Health Research, and the Social Sciences and Humanities Research Council.

The authors acknowledge the tremendous contribution and support of AOF participants and AOF team members. All Our Families is funded through Alberta Innovates Interdisciplinary Team Grant #200700595, the Alberta Children’s Hospital Foundation, the Canadian Institutes of Health Research, and the Social Sciences and Humanities Research Council.

**Avon Longitudinal Study of Parents and Children (ALSPAC)**

We are extremely grateful to all of the families who took part in ALSPAC, the midwives for their help in recruiting them, and the whole ALSPAC team, which includes interviewers, computer and laboratory technicians, clerical workers, research scientists, volunteers, managers, receptionists and nurses.

Core funding for the Avon Longitudinal Study of Parents and Children (ALSPAC) is provided by the UK Medical Research Council and Wellcome (217065/Z/19/Z) and the University of Bristol. A comprehensive list of grants funding is available on the ALSPAC website (http://www.bristol.ac.uk/alspac/external/documents/grant-acknowledgements.pdf). DAL and AK work in a unit that is supported by the University of Bristol and UK Medical Research Council (MC_UU_00011/6) and DAL holds a European Research Council Advanced Grant (ERC grant agreement no 669545) and is a NIHR Senior Investigator (NF-0616-10102). The funders had no role in the design of the study, the collection, analysis, or interpretation of the data; the writing of the manuscript, or the decision to submit the manuscript for publication. The views expressed in this paper are those of the authors and not necessarily those of any funder.

**Amsterdam Born Children and their Development Study (ABCD)**

We are grateful to all participating hospitals, obstetric clinics, and general practitioners for their assistance in implementing the ABCD study and thank all of the women who participated for their cooperation. Core funding of the ABCD-study is provided by the Academic Medical Centre, Amsterdam, the Public Health Services, Amsterdam, and the Dutch Organization for Health Research and Development (ZonMw).

**Babies After SCOPE: Evaluating the Longitudinal Impact on Neurological and Nutritional Endpoints (BASELINE)**

The authors thank the families for their continued support and the Cork BASELINE Birth Cohort Study research team. SCOPE Ireland was supported by the Health Research Board, Ireland (CSA 2007/2). The BASELINE cohort was funded by the National Children’s Research Centre, Dublin, Ireland, and the Food Standards Agency of the United Kingdom (grant no. TO7060).

**Barwon Infant Study (BIS)**

We thank the BIS participants for the generous contribution they have made to this project. We also thank current and past staff for their efforts in recruiting and maintaining the cohort and in obtaining and processing the data and biospecimens.

The establishment work and infrastructure for the BIS was provided by the Murdoch Children’s Research Institute, Deakin University and Barwon Health. Subsequent funding was secured from the National Health and Medical Research Council of Australia, The Jack Brockhoff Foundation, the Scobie Trust, the Shane O’Brien Memorial Asthma Foundation, the Our Women’s Our Children’s Fund Raising Committee Barwon Health, The Shepherd Foundation, the Rotary Club of Geelong, the Ilhan Food Allergy Foundation, GMHBA Limited and the Percy Baxter Charitable Trust, Perpetual Trustees. In-kind support was provided by the Cotton On Foundation and CreativeForce. Research at Murdoch Children’s Research Institute is supported by the Victorian Government's Operational Infrastructure Support Program. This work was also supported by NHMRC Senior Research Fellowships (1064629 to DB; 1045161 to RS) and NHMRC Investigator Grants to DB (1175744).

**Born in Guangzhou Cohort Study (BIGCS)**

We are grateful to the pregnant women who participated in the BIGCS and all obstetric care providers who assisted in the implementation of the study. This work was supported the Department of Science and Technology of Guangdong Province, China (no. 2020B1111170001).

**Clinical review of the Health of 22–33 years old conceived with and without ART (CHART)**

We would like to acknowledge the participants who generously gave their time to the study and the invaluable contribution of Ms. Jane Koleff to the development of the protocol and in the training of all assessors to undertake the clinical assessments.

The CHART study was supported by the Victorian State Government Operational Infrastructure Support and the Australian Government NHMRC IRIISS awarded to the Murdoch Children’s Research Institute, and funded by a National Health & Medical Research Council Project Grant (APP1099641; 2016–2017), Royal Children’s Hospital Research Foundation, Monash IVF Research and Education Foundation, and Reproductive Biology Unit Sperm Fund, Melbourne IVF

**Danish National Birth Cohort (DNBC)**

The authors would like to thank the participants, the first Principal Investigator of DNBC Prof. Jørn Olsen, the scientific managerial team, and DNBC secretariat for being, establishing, developing and consolidating the Danish National Birth Cohort.

The Danish National Birth Cohort was established with a significant grant from the Danish National Research Foundation. Additional support was obtained from the Danish Regional Committees, the Pharmacy Foundation, the Egmont Foundation, the March of Dimes Birth Defects Foundation, the Health Foundation and other minor grants. The DNBC Biobank has been supported by the Novo Nordisk Foundation and the Lundbeck Foundation. Follow-up of mothers and children have been supported by the Danish Medical Research Council (SSVF 0646, 271-08-0839/06-066023, O602-01042B, 0602-02738B), the Lundbeck Foundation (195/04, R100-A9193), The Innovation Fund Denmark 0603-00294B (09-067124), the Nordea Foundation (02-2013-2014), Aarhus Ideas (AU R9-A959-13-S804), University of Copenhagen Strategic Grant (IFSV 2012), and the Danish Council for Independent Research (DFF – 4183-00594 and DFF - 4183-00152).

**Etude de cohorte généraliste, menée en France sur les Déterminants pré et post natals précoces du développement psychomoteur et de la santé de l’Enfant (EDEN)**

The authors thank the cohort participants and the EDEN mother-child study group, whose members are: I. Annesi-Maesano, J.Y. Bernard, J. Botton, M.A. Charles, P. Dargent-Molina, B. de Lauzon-Guillain, P. Ducimetière, M. de Agostini, B. Foliguet, A. Forhan, X. Fritel, A. Germa, V. Goua, R. Hankard, B. Heude, M. Kaminski, B. Larroque†, N. Lelong, J. Lepeule, G. Magnin, L. Marchand, C. Nabet, F Pierre, R. Slama, M.J. Saurel-Cubizolles, M. Schweitzer, O. Thiebaugeorges.

The EDEN study was supported by Foundation for medical research (FRM), National Agency for Research (ANR), National Institute for Research in Public health (IRESP: TGIR cohorte santé 2008 program), French Ministry of Health (DGS), French Ministry of Research, INSERM Bone and Joint Diseases National Research (PRO-A) and Human Nutrition National Research Programs, Paris-Sud University, Nestlé, French National Institute for Population Health Surveillance (InVS), French National Institute for Health Education (INPES), the European Union FP7 programmes (FP7/2007- 2013, HELIX, ESCAPE, ENRIECO, Medall projects), Diabetes National Research Program (through a collaboration with the French Association of Diabetic Patients (AFD)), French Agency for Environmental Health Safety (now ANSES), Mutuelle Générale de l’Education Nationale a complementary health insurance (MGEN), French national agency for food security, French speaking association for the study of diabetes and metabolism (ALFEDIAM).

**Etude Longitudinale Franc¸aise depuis l’Enfance (ELFE)**

The authors are grateful to 1) the former members of the Elfe unit without whom the project would never have started: Henri Léridon, initiator and former Principal Investigator of the project, Stéphanie Vandentorren, Claudine Pirus, and Ando Rakotonirina; 2) the expertise and assistance of members of the unit for support functions, 3) all the researchers who contribute to the projects as members of the Elfe thematic groups and especially their coordinators; 4) all the field research assistants and interviewers; 5) and above all, all the Elfe families who have placed their confidence in us and given up their time to the study.

The Elfe cohort received funding from the National Research Agency Investment for the Future program [ANR-11-EQPX-0038]; French National Institute for Research in Public Health (IRESP TGIR 2009-2001 program); Ministry of Higher Education and Research; Ministry of Environment; Ministry of Health; French Agency for Public Health; Ministry of Culture; and National Family Allowance Fund.

**EU Childhood Obesity Project (CHOP)**

The authors would particularly like to thank all the cohort participants for their generous collaboration. Furthermore, thanks to all persons who designed and conducted the study, entered the data, and participated in the data analysis and who are represented by the European Childhood Obesity Trial Study Group participants: B Koletzko, V Grote, M Totzauer, K Gürlich, P Schwarzfischer, N Aumüller, V Luque, M Zaragoza-Jordana, N Ferré, J Escribano, R Closa-Monasterolo, A Xhonneux, Jean-Paul Langhendries, E Verduci, E Riva, D Gruszfeld.

The CHOP study has been carried out with partial financial support from the Commission of the European Community, specific RTD Programme "Quality of Life and Management of Living Resources", within the Fifth Framework Program (research grants no. QLRT-2001-00389 and QLK1-CT-200230582), the Sixth Framework Program (contract no. 007036), and Seventh Framework Programme (EarlyNutrition; grant agreement no. 289346), the EU H2020 project LIFECYCLE under grant no. 733206 and the European Research Council Advanced Grant META-GROWTH (ERC-2012-AdG – no.322605) and with financial support from Polish Ministry of Science and Higher Education (2571/7.PR/2012/2). This manuscript does not necessarily reflect the views of the Commission and in no way anticipates the future policy in this area. No funding bodies had any role in the study design, data collection and analysis.

**Generation R (Gen R)**

The authors gratefully acknowledge the contribution of participants, research collaborators, general practitioners, hospitals, midwives, and pharmacies in Rotterdam.

The general design of the Generation R Study is made possible by financial support from the Erasmus MC, University Medical Center, Rotterdam, Erasmus University Rotterdam, Netherlands Organization for Health Research and Development (ZonMw), Netherlands Organisation for Scientific Research (NWO), Ministry of Health, Welfare and Sport and Ministry of Youth and Families. This project received funding from the European Union's Horizon 2020 research and innovation programme (LIFECYCLE, grant agreement No 733206, 2016, European Joint Programming Initiative “A Healthy Diet for a Healthy Life” (JPI HDHL, EndObesity project, ZonMW the Netherlands no. 529051026). RG received funding of the Dutch Heart Foundation (grant number 2017T013), the Dutch Diabetes Foundation (grant number 2017.81.002), and the Netherlands Organization for Health Research and Development (NWO, ZonMW, grant number 543003109). The study sponsors had no role in the study design, data analysis, interpretation of data, or writing of this report.

**Generation XXI (G2I)**

G21 was funded by Programa Operacional de Saúde – Saúde XXI, Quadro Comunitário de Apoio III and Administração Regional de Saúde Norte (Regional Department of Ministry of Health) and by Foundation for Science and Technology – FCT (UIDB/04750/2020 - Unidade de Investigação em Epidemiologia (EPIUnit), Instituto de Saúde Pública da Universidade do Porto). The funders had no role in study design, data collection and analysis, interpretation of data, or writing of this report.

**Growing Up in Ireland Infant Cohort (GUI)**

The authors sincerely thank the thousands of Irish families that contribute to the Growing Up in Ireland Project. None of this would be possible without your time and effort.

Growing Up in Ireland (GUI) is funded by the Department of Children, Equality, Disability, Integration and Youth (DCEDIY). It is being carried out by a consortium of researchers led by the Economic and Social Research Institute (ESRI) and Trinity College Dublin (TCD). GUI is managed by DCEDIY in association with the Central Statistics Office (CSO). Results in this report are based on analyses of data from Research Microdata Files provided by the Central Statistics Office (CSO). Neither the CSO nor DCEDIY take any responsibility for the views expressed or the outputs generated from these analyses.

**Growing Up in New Zealand (GUiNZ)**

We thank the participating families of the *Growing Up in New Zealand* cohort study who have given their time and shared the information that allowed us to conduct this research. The *Growing Up in New Zealand* study has been funded by the New Zealand Ministries of Social Development, Health, Education and Justice; the former Ministry of Science Innovation and the former Department of Labour (now both part of the Ministry of Business, Innovation and Employment); the former Ministry of Pacific Island Affairs (now the Ministry for Pacific Peoples); the former Ministry of Women’s Affairs (now the Ministry for Women); the Department of Corrections; the Families Commission and the former Social Policy Evaluation and Research Unit; Te Puni Kokiri; New Zealand Police; Sport New Zealand; Housing New Zealand Corporation; and the former Mental Health Commission (now part of the Office of the Health and Disability Commissioner); The University of Auckland and Auckland UniServices Limited. Other support for the study has been provided by the Health Research Council of New Zealand, Statistics New Zealand, the Office of the Children’s Commissioner and the Office of Ethnic Affairs (now the Office of Ethnic Communities).

**Growing up in Singapore Towards healthy Outcomes (GUSTO)**

We thank the GUSTO study group and all clinical and home-visit staff involved. The voluntary participation of all participants is greatly appreciated.

The GUSTO study group includes. Allan Sheppard,Amutha Chinnadurai, Anne Ferguson-Smith, Anne Eng Neo Goh, Arijit Biswas, Audrey Chia, Birit Leutscher-Broekman, Borys Shuter, Shirong Cai, Cheryl Ngo, Chai Kiat Chng, Shang Chee Chong, Christiani Jeyakumar Henry, Mei Chien Chua, Cornelia Yin Ing Chee, Yam Thiam Daniel Goh, Dennis Bier, Chun Ming Ding, Doris Fok, Eric Andrew Finkelstein, Fabian Kok Peng Yap, George Seow Heong Yeo, Wee Meng Han, Helen Chen, Hugo P S Van Bever, Hazel Inskip, Iliana Magiati, Inez Bik Yun Wong, Jeevesh Kapur, Jenny L Richmond, Jerry Kok Yen Chan, Joshua J Gooley, Krishnamoorthy Niduvaje, Bee Wah Lee, Yung Seng Lee, Leher Singh, Sok Bee Lim, Lourdes Mary Daniel, Seong Feei Loh, Yen-Ling Low, Pei-Chi Lynette Shek, Marielle Fortier, Mark Hanson, Mary Foong-Fong Chong, Michael Meaney, Susan Morton, Wei Wei Pang, Pratibha Agarwal, Anqi Qiu, Boon Long Quah, Rob M van Dam, David Stringer, Salome Antonette Rebello, Wing Chee So, Chin-Ying Hsu, Lin Lin Su, Jenny Tang, Kok Hian Tan, Soek Hui Tan, Oon Hoe Teoh, Victor Samuel Rajadurai, PC Wong and Sudhakar K Venkatesh

**Healthy Growth Study (HGS)**

The HGS was co-funded by the European Union (European Social Fund – ESF) and Greek national funds through the Operational Program "Education and Lifelong Learning" of the National Strategic Reference Framework (NSRF) - Research Funding Program: Heracleitus II. Investing in knowledge society through the European Social Fund.

**Italian Twin register (ITR)**

The authors thank all twins enrolled in the ITR for study participation. ITR has been funded by the Italian Ministry of Health (MINSAN) (D.L. 502/92, finalised research and by the GenomEUtwin Project (European Union Contract no. QLG2-CT-2002-01254)

**Millenium Cohort Study (MCS)**

We are grateful to the Centre for Longitudinal Studies (CLS), UCL Social Research Institute, for the use of these data and to the UK Data Service for making them available, and to the children and families who take part in the study. Neither CLS nor the UK Data Service bear any responsibility for the analysis or interpretation of these data.

**Multiple Birth Cohort Study (MUBICOS)**

The authors thank all MUBICOS families for participation. MUBICOS was partly funded by Chiesi Onlus Foundation.

**Nascita e INFanzia: gli Effetti dell'Ambiente (NINFEA)**

The authors thank all families participating in the NINFEA cohort.

The NINFEA cohort was initially funded by the Compagnia SanPaolo Foundation and the Piedmont Region. It received funding from European projects: CHICOS (FP7 grant number HEALTH-FP7-2009-241604, LifeCycle (H2020 grant number 733206), ATHLETE (H2020 grant number 874583).

**Norwegian Mother, Father and Child Cohort Study (MoBa)**

The authors are grateful to all the participating families in Norway who take part in this on-going cohort study.

The Norwegian Mother, Father and Child Cohort Study is supported by the Norwegian Ministry of Health and Care Services and the Ministry of Education and Research.

The study was partly funded by the Norwegian Research Council’s Centres of Excellence Funding Scheme, no 262700. MCM is funded by the European Research Council (ERC) under the European Union’s Horizon 2020 research and innovation programme (grant agreement No 947684)

Data from the Norwegian Mother, Father and Child Cohort Study and the Medical Birth Registry of Norway used in this study are managed by the national health register holders in Norway (Norwegian Institute of public health) and can be made available to researchers, provided approval from the Regional Committees for Medical and Health Research Ethics (REC), compliance with the EU General Data Protection Regulation (GDPR) and approval from the data owners. The consent given by the participants does not open for storage of data on an individual level in repositories or journals. Researchers who want access to data sets for replication should apply through helsedata.no. Access to data sets requires approval from The Regional Committee for Medical and Health Research Ethics in Norway and an agreement with MoBa.

**Piccolipiù**

The authors thank all the families who took part in the study, and the Piccolipiù research group.

The study was funded by the Italian National Centre for Disease Prevention and Control (CCM grant 2010), by the Italian Ministry of Health (art 12 and 12bis Dl .gs. vo 502/92).

**Prospective Study on Infancy in Italy (GASPII)**

The authors are grateful to all the participating families in Rome who take part in this on-going cohort study, and to all the field workers and interviewers of the project. The study was funded by Italian Ministry of Health.

**Southampton Women's Survey (SWS)**

The authors are grateful to the women of Southampton who gave their time to take part in the Southampton Women’s Survey and to the research nurses and other staff who collected and processed the data.

The SWS is supported by grants from the Medical Research Council, National Institute for Health Research Southampton Biomedical Research Centre, British Heart Foundation, UK Food Standards Agency, British Lung Foundation, Versus Arthritis, University of Southampton and University Hospital Southampton National Health Service Foundation Trust, and the European Union’s Seventh Framework Programme (FP7/2007-2013), project Early Nutrition (grant 289346) and from the European Union's Horizon 2020 research and innovation programme (LIFECYCLE, grant agreement No 733206). Study participants were drawn from a cohort study funded by the Medical Research Council and the Dunhill Medical Trust.

**The Trøndelag Health Study (HUNT)**

The Trøndelag Health Study (HUNT) is a collaboration between HUNT Research Centre (Faculty of Medicine and Health Sciences, NTNU, Norwegian University of Science and Technology), Trøndelag County Council, Central Norway Regional Health Authority, and the Norwegian Institute of Public Health.
